# A prognostic molecular signature of hepatic steatosis is spatially heterogeneous and dynamic in human liver

**DOI:** 10.1101/2024.01.26.24301828

**Authors:** Andrew S. Perry, Niran Hadad, Emeli Chatterjee, Maria Jimenez Ramos, Eric Farber-Eger, Rashedeh Roshani, Lindsay K. Stolze, Shilin Zhao, Liesbet Martens, Timothy J. Kendall, Tinne Thone, Kaushik Amancherla, Samuel Bailin, Curtis L. Gabriel, John Koethe, J. Jeffrey Carr, James Greg Terry, Jane Freedman, Kahraman Tanriverdi, Eric Alsop, Kendall Van Keuren-Jensen, John F.K. Sauld, Gautam Mahajan, Sadiya Khan, Laura Colangelo, Matthew Nayor, Susan Fisher-Hoch, Joseph McCormick, Kari E. North, Jennifer Below, Quinn Wells, Dale Abel, Ravi Kalhan, Charlotte Scott, Martin Guilliams, Jonathan A. Fallowfield, Nicholas E. Banovich, Saumya Das, Ravi Shah

## Abstract

Metabolic dysfunction-associated steatotic liver disease (MASLD) prevalence is increasing in parallel with an obesity pandemic, calling for novel strategies for prevention and treatment. We defined a circulating proteome of human MASLD across ≈7000 proteins in ≈5000 individuals from diverse, at-risk populations across the metabolic health spectrum, demonstrating reproducible diagnostic performance and specifying both known and novel metabolic pathways relevant to MASLD (central carbon and amino acid metabolism, hepatocyte regeneration, inflammation, fibrosis, insulin sensitivity). A parsimonious proteomic signature of MASLD was associated with a protection from MASLD and its related multi-system metabolic consequences in >26000 free-living individuals, with an additive effect to polygenic risk. The MASLD proteome was encoded by genes that demonstrated transcriptional enrichment in liver, with spatial transcriptional activity in areas of steatosis in human liver biopsy and dynamicity for select targets in human liver across stages of steatosis. We replicated several top relations from proteomics and spatial tissue transcriptomics in a humanized “liver-on-a-chip” model of MASLD, highlighting the power of a full translational approach to discovery in MASLD. Collectively, these results underscore utility of blood-based proteomics as a dynamic “liquid biopsy” of human liver relevant to clinical biomarker and mechanistic applications.

## INTRODUCTION

Metabolic dysfunction-associated steatotic liver disease (MASLD) is present in >30% of individuals worldwide^1^ and has emerged as a predominant driver of end-stage liver disease and an important contributor to a range of other chronic, life-limiting illnesses (e.g., cancer, cardiovascular disease, renal dysfunction)^2^. The heterogeneous progression from MASLD to chronic steatohepatitis—and limited effectiveness of therapies that interrupt this process—have fueled efforts to identify at-risk populations early for prevention. While large studies of genetic propensity for MASLD have yielded promising targets (e.g., *PNPLA3*^3^), genetic variation alone may not capture dynamic behavioral and environmental contributions (diet, obesity, diabetes, etc.) that may be reflected in molecular states that precede MASLD. Circulating proteomics^4–6^ and hepatic tissue profiles^7^ have been at the forefront of resolving these limitations in MASLD, though limited by lack of integration of circulating “omics” and liver characterization or reliance on invasive biopsy-guided approaches. In response, a few recent studies have begun to integrate liver phenotypes and circulating proteomics in humans with some promising results, albeit in small cohorts with available liver tissue and in targeted populations with specific exposures (e.g., alcohol)^8,9^. Integrative studies that link the circulating proteome to dynamic hepatic tissue states across the MASLD spectrum (at single cell and spatial resolution) and large clinical longitudinal studies to assess clinical susceptibility and consequences of MASLD will provide the broad translational landscape necessary to simultaneously specify diagnostic and prognostic biomarkers and prioritize dynamic targets for mechanistic inquiry.

We leveraged three large prospective observational cohorts (N=4996) across a broad spectrum of metabolic risk with complementary non-invasive measures of hepatic steatosis to identify a circulating proteome of MASLD. Single-protein and multi-protein signatures of MASLD were associated with imaging-defined MASLD across a spectrum of metabolic risk, with clinical diagnostic performance beyond known MASLD risk factors. Identified proteins specified pathways of central carbon and amino acid metabolism, hepatocyte regeneration, inflammation, fibrosis, and insulin sensitivity. Moreover, this multivariable signature of MASLD was associated with incident non-alcoholic chronic liver disease, diabetes, and a host of metabolic conditions in >26000 participants over nearly a decade of follow-up in the UK Biobank, with additivity over polygenic risk estimates and prognostic reliability down to a clinically translatable 21-protein panel. We observed very strong enrichment of the MASLD-associated proteome in human liver at a transcriptional level, as well as spatial transcriptional localization to areas of histologically defined steatosis in human liver in early MASLD. Expression of key targets appeared predominantly in hepatocytes, macrophages, and fibroblasts and several targets’ (prioritized via spatial transcription and proteomics) expression correlated with progression of steatosis in >600 human livers and in a humanized “liver-on-a-chip” model of early MASLD. Collectively, these results inform fundamental connections between a blood-based proteomic signature of MASLD as a “liquid biopsy” of potentially dynamic states in human liver, connecting clinical biomarker discovery and hepatic biology in MASLD.

## RESULTS

### General flow of study and characteristics of study samples

Our study consisted of five integrated steps (**Figure 1**; details in **Methods**), specifically (1) identification and validation of a “MASLD proteome” (across 4996 participants across three prospective observational studies: Coronary Artery Risk Development in Young Adults [CARDIA], UK Biobank, Cameron County Hispanic Cohort [CCHC]); (2) characterization of tissue origin and implicated molecular pathways; (3) relation of this proteome to MASLD-relevant clinical outcomes, including complementarity with human genetics (across 26421 participants in UK Biobank over a median 13.7 years of follow-up); (4) examining expression of genes encoding the MASLD proteome in human liver across stages of MASLD (bulk RNA-seq in human liver; SteatoSITE, N=523 biopsies); (5) specifying cell and spatially resolved expression of these genes in human liver with and without MASLD (scRNA-seq, N=19; spatial transcriptomics, N=4).

**Figure 1:**
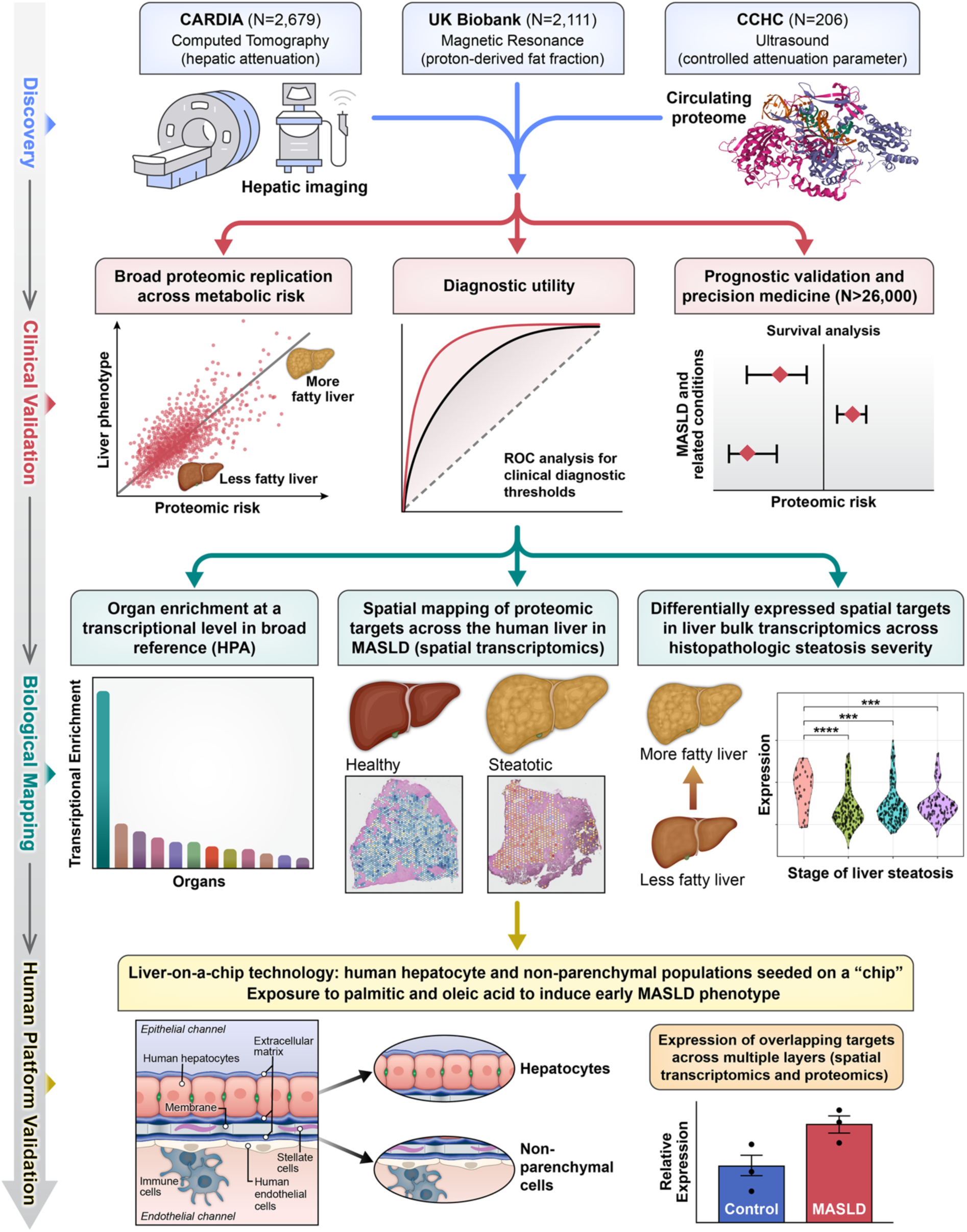
Study diagram: Study design.

The characteristics of the study samples are shown in **Supplementary Tables 1-3**. Overall, our study included 2679 CARDIA study participants after excluding participants with other potential causes for hepatic steatosis (>14 alcoholic drinks/week, hepatitis C, cirrhosis, HIV, and use of amiodarone, valproic acid, methotrexate, tamoxifen, or diltiazem)^10^. CARDIA participants were randomly split into derivation (N=1876) and validation (N=803) subsamples, with an overall median age of 51 years, 57% female, and 47% Black individuals (**Supplemental Table 1**). Participants were predominantly overweight and obese (median body mass index [BMI] 29 kg/m^2^), with low alcohol use (median 1 drink/week) and 15% had diabetes. The study population from UK Biobank included 26421 participants that represented a broader age range (25^th^ to 75^th^ percentile: 50-64 years), 54% were women, and participants were predominantly White (94%) and overweight (median 27kg/m^2^) with a lower prevalence of diabetes (5.8%) and greater alcohol intake (43% reporting ≥ 3 times a week). A total of 2111 UK Biobank participants had MRI measures of hepatic steatosis (**Supplemental Table 2**). Participants from the Cameron County Hispanic cohort (CCHC; N=206) had similar age (25^th^ to 75^th^ percentile: 46-66 years) and sex (66% female) distributions, composed entirely of White Hispanics, with greater obesity (median BMI 31 kg/m^2^) and prevalence of diabetes (32%), with 49% reporting no alcohol use (**Supplemental Table 3**).

### Circulating proteomics of MASLD identify broad canonical pathways of human metabolism with predominant gene expression in the liver

Across 2679 participants in CARDIA with SomaScan 7k proteomics, we identified 237 unique proteins (259 SomaScan aptamers) associated with liver attenuation on computed tomography (lower liver attenuation ∼ more hepatic steatosis) across both derivation and validation subsamples (adjusted for age, sex, race, BMI; **Figure 2A-B**; full regression estimates in **Supplementary Data File SD02**). Regression estimates were robust to multivariable adjustment, including metabolic risk factors, renal function, physical activity, and alcoholic drinks per week (relation of regression estimates across adjustments: Spearman ρ=0.95; P < 2.2×10^−16^; **Supplemental Figure 1**). We observed significant enrichment of genes encoding the MASLD proteome in the liver (**Figure 2C**), specifying broad pathways implicated in central metabolic processes (e.g., carbon, pyruvate, amino acid, carbohydrate metabolism) and fibrosis (**Figure 2D**), including known and emerging mechanisms of MASLD, namely amino acid metabolism (*ACY1*^6,11^, *FAH*^12^), alcohol processing (e.g., *ADH1A*^13,14^), fructose catabolism (*ALDOB, SORD*^15^), bile acid and steroid metabolism (*AKR1D1*^16^, *AKR1C4*^17,18^), gluconeogenesis (*FBP1*^19^), and multi-substrate detoxification, intermediary metabolism, and fibrosis (*GSTA1*^20^*, ASL*^21^*, UGDH*^22^), among others. To identify potential central mediators of MASLD, we next conducted an interaction (hub gene) analysis including 235 genes (of the 237 unique genes), with nodes identifying genes with central relevance to MASLD (**Figure 2E**). Identified nodes included pathogenic mediators of hepatocyte regeneration and fibrosis regulation (*EGFR*^23^, *IGF-1*^24^), apoptosis regulation (*MET*^25^), inflammatory mediators (*CXCL2*^26^, *CRP*, *SERPINE1*), extracellular matrix responses to hepatic injury (*VTN*^27^, *ACAN*^28^), glycogen metabolism (*PYGL*^29^), and mitochondrial pyruvate metabolism (*PKLR*^30^, *PC*^31^), among several other canonical markers of insulin sensitivity and adiposity (*ADIPOQ*, *INS*). These results suggested a predominant hepatic origin for the circulating MASLD proteome, implicating canonical metabolic-inflammatory-fibrotic pathways in liver degeneration.

**Figure 2.**
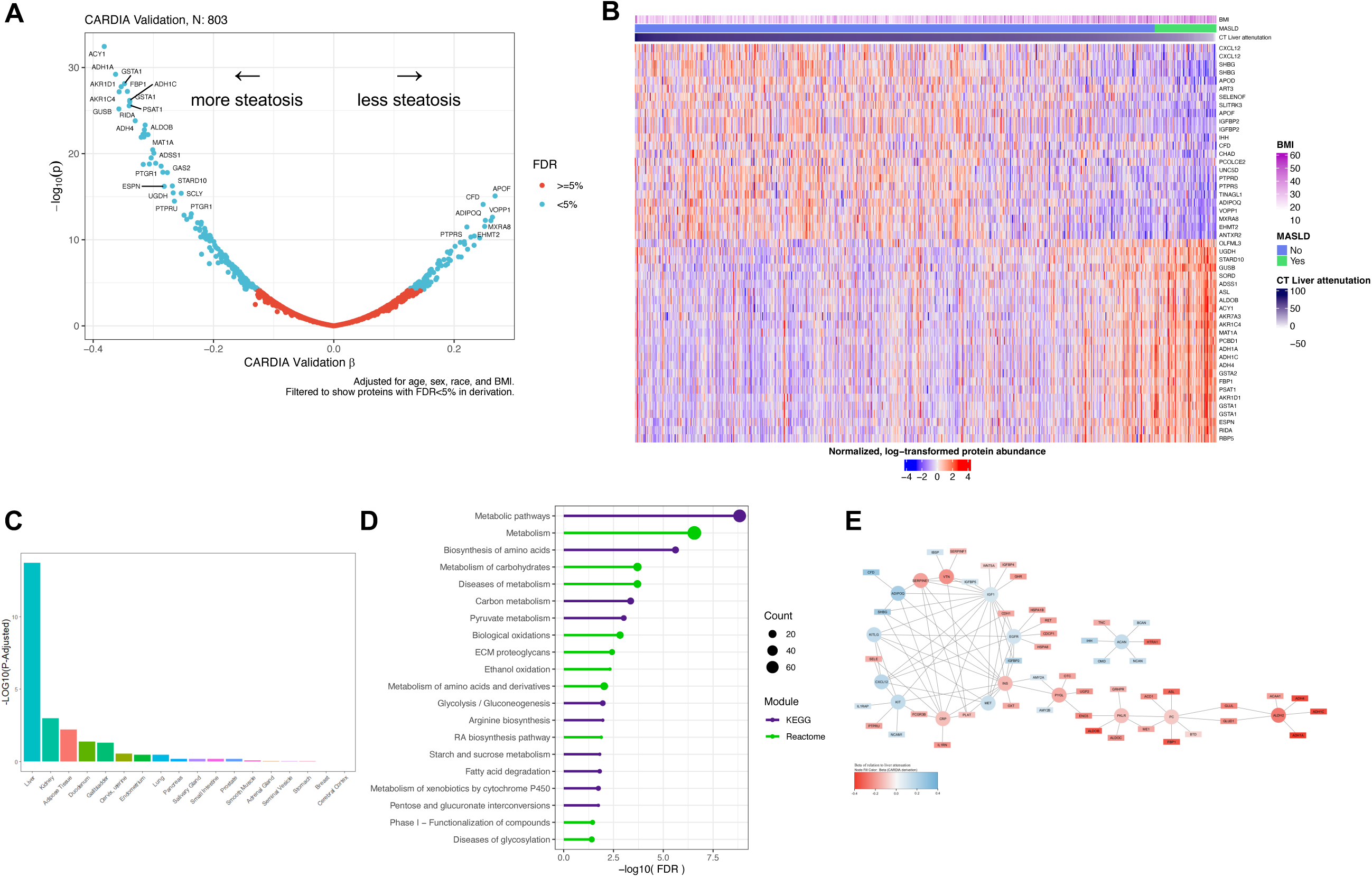
Proteins related to hepatic steatosis are primarily expressed in the liver and identify pathways of metabolism. (**A**) Volcano plot of proteins associated with hepatic steatosis after adjustment for age, sex, race, and BMI. For visualization, proteins with an FDR<5% in CARDIA derivation subsample are visualized with the beta coefficient and p values presented coming from models using the CARDIA validation subsample. (**B**) Heatmap of the top 25 positively associated and top 25 negatively associated proteins with hepatic steatosis in the CARDIA validation sample. (**C**) Tissue expression analysis of the proteins related to hepatic steatosis in CARDIA using the full SomaScan 7k platform as the background, demonstrated enrichment of proteins expressed in the liver. (**D**) KEGG and Reactome pathway analysis. (**E**) Hub gene analysis of significant proteins associated with liver attenuation showing the hub genes (≥5 connections; circles) and all proteins with high confidence connections to the hub genes (rectangles).

### Integrated proteomics identifies a diagnostic biomarker of MASLD with wide replication across metabolic risk states

Penalized regression (LASSO) generated a 336-aptamer model for liver attenuation adjusted for age, sex, race, and BMI (hereafter referred to as “MASLD score”). The MASLD score correlated with liver attenuation in both derivation and validation subsamples within CARDIA (Spearman ρ=0.69 and 0.56, respectively; **Figure 3A**; model coefficients in **Supplementary Data File SD04**), differed across clinical thresholds for MASLD (**Figures 3B, 3D**), with retention of correlations using the top 21 aptamers from the MASLD score (**Supplemental Figure 3A**). Addition of the MASLD score to standard clinical risk (age, sex, race, BMI, alcoholic drinks per week, aspartate aminotransferase [AST; aptamer based], alanine aminotransferase [ALT; aptamer based], hemoglobin A1c) markedly improved MASLD discrimination (C-index 0.84 [95% CI 0.80-0.88] to 0.94 [95% CI 0.92-0.96], P=4.3×10^−7^; **Figure 3F**). This discriminative performance was maintained using the top 21 aptamers from the MASLD score (21 chosen given the limit of current absolute proteomic detection [Olink] for clinical translation). Of note, despite the well described association between obesity and MASLD, we only observed a modest correlation between BMI and the protein score (Spearman ρ=-0.27, P<2.2×10^−16^), with smaller effects by age, sex, race, and alcohol use (**Supplemental Figure 2)**.

**Figure 3:**
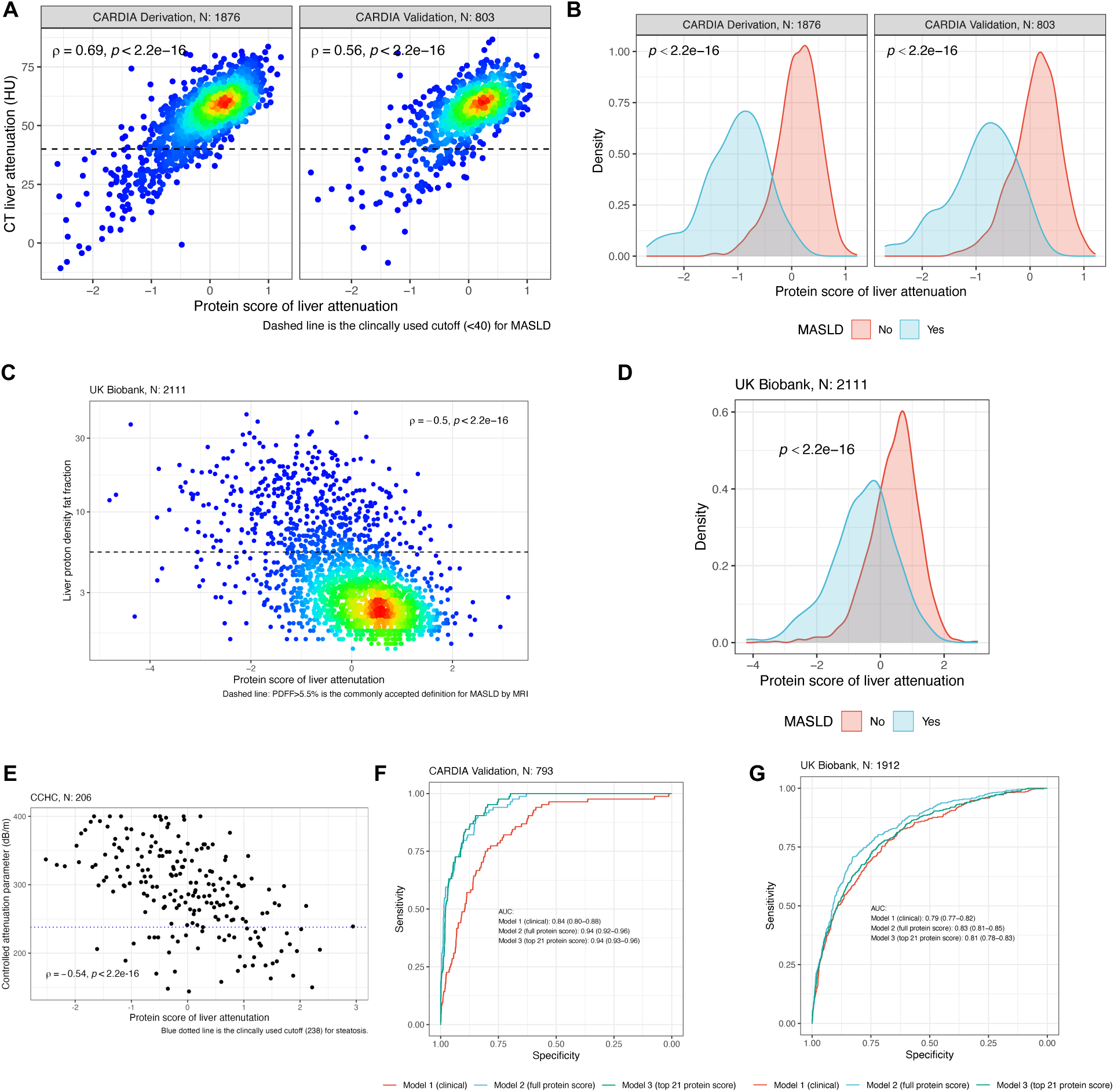
Development of a proteomic score of MASLD and its utility as a diagnostic test. (**A**) A protein score of liver attenuation by CT (less attenuation ∼ more steatosis) demonstrated moderate correlation with the parent variable in both CARDIA derivation and validation samples. (**B**) The protein score distinguishes between MASLD and non-MASLD populations in CARDIA. (**C**) Replication of the association between a protein score of liver attenuation and MRI-based measure of hepatic steatosis (proton density fat fraction: higher ∼ more steatosis, opposite directionality as with CT based liver attenuation) in UK Biobank. (**D**) The protein score distinguishes between MASLD and non-MASLD populations in UK Biobank. (**E**) The protein score is related to controlled attenuation parameter (higher value ∼ more steatosis) in CCHC. (**F**) Receiver operator curve analysis in the CARDIA validation sample comparing a clinical model of MASLD (age, sex, race, BMI, drinks/week, AST, ALT, A1c) to models including a protein score of liver attenuation. (**G**) ROC analysis in UK Biobank.

For external rigor of our approach, we next replicated the MASLD score with hepatic steatosis in >2000 participants from 2 different studies with distinct approaches to MASLD quantification (CCHC, ultrasound; MRI, UK Biobank). As CCHC and UK Biobank used Olink proteomics platforms, recalibration of the MASLD score was performed (see **Methods**). Of note, CT liver attenuation (the measure in CARDIA) is an opposite directionality relative to ultrasound or MRI (higher attenuation ∼ lower steatosis ∼ lower MRI or ultrasound measure). In 2111 UK Biobank participants, our results were largely similar, including (1) a similar relation between the recalibrated MASLD score and MRI-determined proton-derived fat fraction (Spearman ρ=-0.5, P<5.9×10^-135^); **Figure 3C, Supplemental Figure 3B**); (2) improvement in MASLD discrimination above clinical models (defined as PDFF>5.5%, a clinically accepted threshold^32^; C-statistic 0.79 [0.77-0.82] to 0.83 [0.81-0.85], P=5.2×10^−8^, **Figure 3G**); (3) similar associations with age, sex, race, BMI, and alcohol use (**Supplemental Figure 2**). Similarly, in CCHC—despite at far higher metabolic risk with far more prevalent MASLD (**Supplemental Table 3**)—we found a magnitude of correlation consistent with other cohorts for controlled attenuation parameter (an ultrasound-based measure of steatosis; Spearman ρ=-0.54, P<2.2×10^−16^; **Figure 3E**).

Recognizing the importance of adiposity on the risk of MASLD, we interrogated whether the relationship between the MASLD score and hepatic steatosis was modified by overweight/obesity status. In the CARDIA validation subsample (N=803), we observed a stronger correlation among participants with obesity (BMI≥30 kg/m^2^; Spearman ρ=0.65) than participants with normal/overweight BMI (<30 kg/m^2^; Spearman ρ=0.41; **Supplemental Figure 4**). In a linear model for the outcome of hepatic steatosis (which was CT assessed liver attenuation in CARDIA), we observed a statistically significant interaction between the MASLD score and BMI wherein the magnitude of the relationship between the MASLD score and CT liver attenuation increases with greater BMI (interaction β=7.5, P=1.4×10^−8^). In conjunction with the observed relationship in CCHC (a cohort with elevated metabolic risk), these findings support the relevance of the MASLD score in at-risk populations.

### A MASLD proteomic score identifies individuals at long-term chronic non-alcoholic liver disease risk, MASLD-related diseases, and survival in over 26,000 people, additive to polygenic risk

In 26421 participants in UK Biobank (median follow-up for mortality 13.7 years, 25^th^-75^th^ percentile 13.0-14.5 years), we found a broad relation of the MASLD score with MASLD and MASLD-relevant clinical outcomes (**Figure 4A**). In addition to all-cause and cause-specific mortality (specifically cancer), we observed very strong relations between the MASLD score and chronic non-alcoholic liver diseases (an electronic health record surrogate of MASLD; adjusted HR 0.59, 95% 0.52-0.67; P=7.0×10^−16^) and diabetes (adjusted HR 0.50, 95% 0.47-0.54; P=1.7×10^−86^; adjustments in **Methods**). In sensitivity analyses, these associations were robust to additional adjustment for AST, ALT, and hemoglobin A1c (chronic non-alcoholic liver disease HR 0.63, diabetes HR 0.53; **Supplemental Data File SD06**) and were retained down to a 21-protein Olink panel (chronic nonalcoholic liver disease HR 0.64, diabetes HR 0.57; **Figure 4C**). In addition to association with clinical outcomes, addition of the MASLD score provided marked improvements in discrimination and net reclassification above clinical models (including age, sex, race, BMI, systolic blood pressure, diabetes [removed for models of diabetes], Townsend Deprivation Index, smoking, alcohol use, and low-density lipoprotein) for both diabetes (C-statistic 0.79 vs. 0.84, P=6.6×10^−20^) and chronic non-alcoholic liver disease (C-statistic 0.69 vs. 0.74, P=1.1×10^−5^; **Figure 4A**). In a sensitivity analysis for incident chronic non-alcoholic liver disease we further adjusted for AST, ALT and A1c and still observed a significant improvement in model performance (C-statistic 0.72 vs 0.74, P=0.009). Given the widespread use of polygenic liability of diabetes for clinical risk prediction^33^, we tested the relative effect of the MASLD score and a diabetes polygenic risk score (PRS), demonstrating a largely additive effect with minimal interaction (weak PRS-by-proteomic interaction beta = 0.07, P=0.001; model predicted hazard ratios presented in **Figure 4B**). These results suggested a strong proteomic liability to incident MASLD and MASLD-related disorders, with significant addition to polygenic risk estimates for risk stratification.

**Figure 4:**
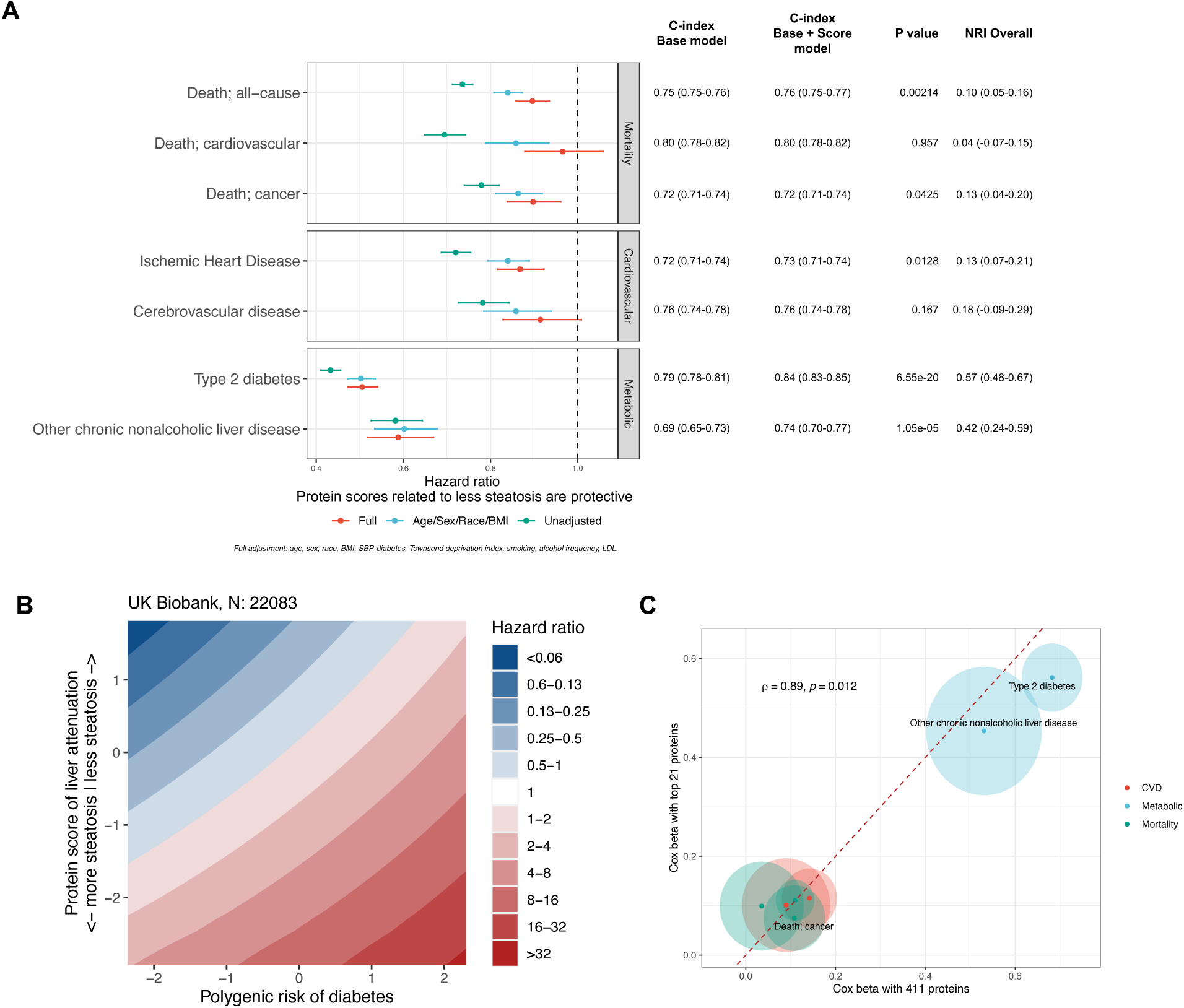
Protein score of liver attenuation is strongly associated with incident metabolic disease and additive to genetic risk. (**A**) Forest plot of associations with clinical outcomes in UK Biobank along with C-index comparisons of models with and without the protein score (see **Supplemental Data File SD06**). P values reported are for comparisons of C-indices. (**B**) We observed a weak interaction between the protein score of liver attenuation and polygenic risk for incident diabetes suggesting the effects of both are largely complementary and additive. Hazard ratios presented are from model predicted estimates. (**C**) Cox regression models using a clinically translatable 21-protein panel provides similar prognostication as the full protein score (see **Supplemental Data File SD06**). The shaded ellipses represent the standard error of the beta coefficient.

### The MASLD proteome exhibits directionally consistent, cell-specific, and spatially localized expression in areas of steatosis in early MASLD with dynamicity during MASLD progression

To elucidate the spatial and cellular organization of prioritized targets identified in proteomic analyses, we conducted a comprehensive mapping of proteins implicated in regression in CARDIA to human liver tissue in early MASLD. Leveraging published single-cell and single-nuclear RNA sequencing (scRNA-seq) and spatial transcriptomics^34^ (N=21; 62% women, mean age 59 years, mean BMI 32 kg/m^2^), we investigated the expression profiles of 198 MASLD-associated protein-gene overlaps represented in both the scRNA-seq and spatial transcriptomics datasets using individual gene expression and a composite expression score (**Figure 5**, see **Methods**) to facilitate the broad identification of cell types and spatial distribution of targets prioritized by the MASLD proteome within the human liver. This analysis showed a predominant cell-specific expression pattern of implicated targets, primarily observed in hepatocytes. However, a subset of targets implicated by the MASLD proteome show heterogenous expression pattens across cell, including fibroblasts, cholangiocytes, endothelial cells, and immune cells (**Figure 5A** and **Supplemental Figure 5A**). Employing a composite expression score, we showed higher gene activity of implicated targets within steatotic tissue and in the mid-central liver zonation. These zones were previously shown to correspond with higher hepatocyte expression signature using the same dataset (**Figure 5B-F, Supplemental Figure 5B)**^34^.

**Figure 5:**
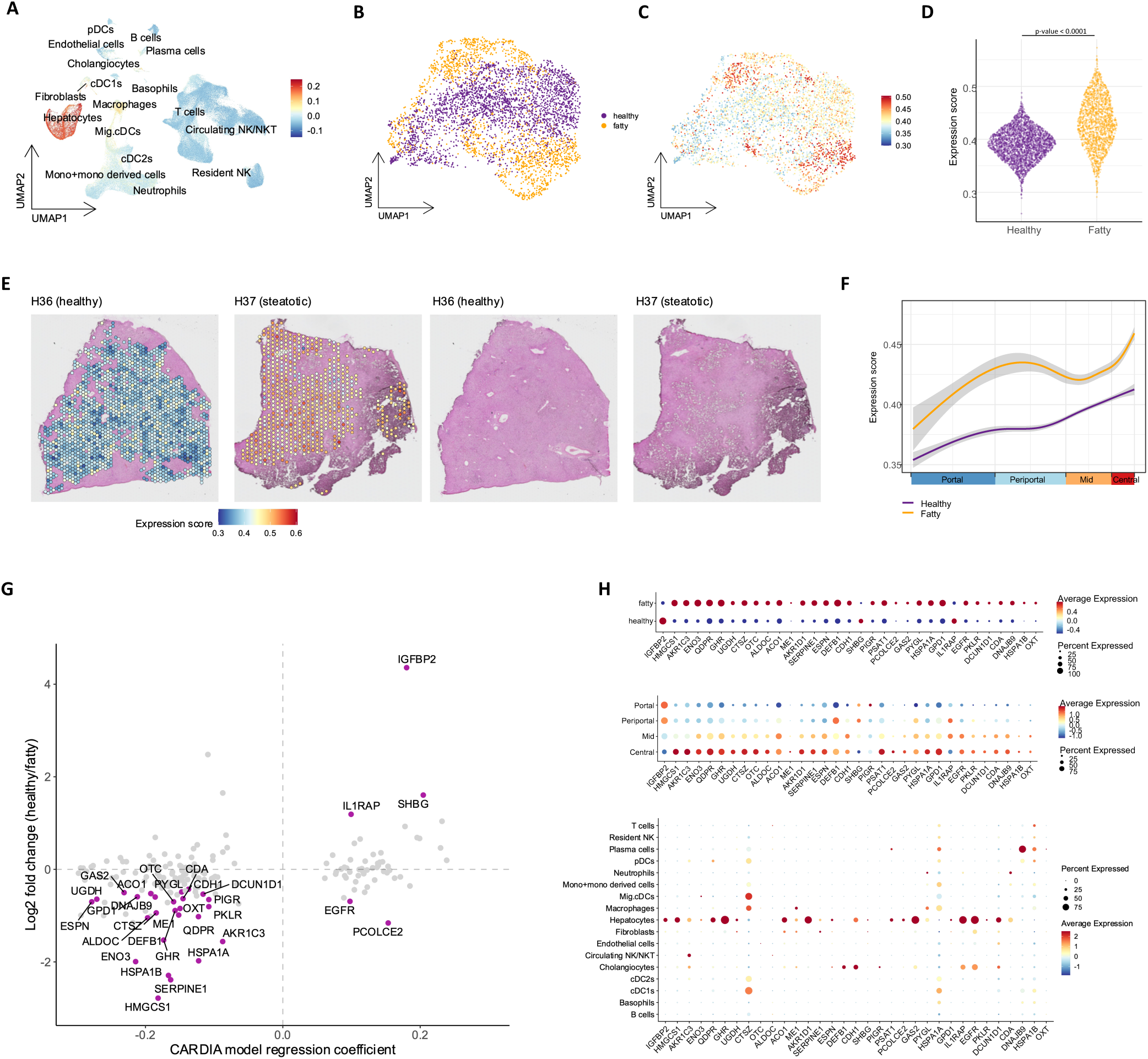
Single nuclear and spatial transcriptional architecture of the circulating MASLD proteome. (**A**) Uniform manifold approximation projection (UMAP) of single nuclear RNA sequencing in steatotic and healthy liver, colored by a composite expression score (derived from gene expression of implicated target proteins, see **Methods**). (**B**, **C**) UMAP of spatial transcriptomic data (Visium) in the liver colored by liver pathology diagnosis (healthy vs. fatty, **B**) and the composite expression score (**C**). (**D**) Violin plot comparing composite expression score across Visium spots by fatty vs. healthy state (Wilcoxon rank-sum). (**E**) Representative images of healthy and steatotic hematoxylin-eosin-stained liver tissue overlaid with Visium spots, colored by composite expression score, demonstrating increased activity of implicated targets in steatotic regions. All sections presented in our parent manuscript are shown in **Supplemental Figure 4**. (**F**) Composite expression across liver zonation groups. (**G**) Differential expression of implicated targets between healthy and fatty liver (Visium) versus circulating proteomic regression coefficient. A more positive proteomic coefficient indicates less liver fat, and a more positive log2 fold-change indicates greater expression of a given transcript in healthy (non-steatotic) liver. Highlighted in purple are targets that were considered as differentially expressed using spatial data (satisfied adjusted p-value < 0.05 & absolute log2 fold-change > 0.25). (**H**) Gene expression of significantly different implicated targets between healthy and steatotic regions (Visium), liver zonation (Visium) and across cell types (single-cell RNA sequencing).

We subsequently assessed whether implicated targets identified by the CARDIA model exhibit differential expressed in single-cell data. To account for the confounding effects of cell isolation technique on the liver cell atlas cell types, we specifically utilized the spatial transcriptomics dataset for this analysis. Of the 198 genes encoding implicated proteins expressed in the spatial transcriptomics Visium dataset, 33 were differentially expressed between healthy and steatotic tissue (minimum expression > 10% of spots; adjusted P < 0.05; |log2 fold-change| > 0.25 & |minimum % expressed spot difference| > 10%; **Figure 5H**). Of the 33 genes, 30 were upregulated in steatotic tissue and only 3 genes (*IGFBP2*, *IL1RAP* and *SHBG*) were downregulated. These genes exhibited distinct expression patterns in human liver tissue with and without steatosis (refer to **Figure 5G-H**, select targets shown in **Supplemental Figure 5C-D**. Moreover, we observed high biological concordance between their change in expression and the clinical effect estimate of the <u>circulating protein</u> corresponding to this gene with liver attenuation in CARDIA (**Figure 5G**). One notable example with high effect size is *IGFBP2*—dynamic during metabolic intervention^35^ and regeneration^36^ with liver-enriched expression^37^—which was increased in healthy versus steatotic cell populations (transcriptional level, **Supplemental Figure 5D**) and associated with lesser steatosis in CARDIA (population proteomic level), consistent with smaller reports^38^. Most observed population association-tissue concordance consisted of metabolic genes up-regulated in steatotic liver that displayed greater circulating protein abundance in individuals with steatosis (**Figure 5G-H**), many of which had established mechanistic relevance in model systems of MASLD and its progression (e.g., *ENO3* and ferroptosis^39^; *UGDH* and fibrosis/redox status^22^; *CTSZ* and epithelial-mesenchymal transition^40^; *CDH1* and lipogenesis^41^; *CDH1* and *PPAR*/*PGC1a* signaling^42^; among others).

Next, we matched these 33 differentially expressed genes to bulk transcriptomic data across NASH-CRN defined stages of hepatic steatosis to investigate potential dynamicity across individuals with increasing severity of histopathologic phenotype (**Figure 6**). Of 523 SteatoSITE participants with biopsy samples, there were 489 with MASLD (54% women, mean age 52 years, mean BMI 32 kg/m^2^, 75% diabetes) compared to 34 control samples. Of the 33 genes passed forward for assessment in bulk transcriptomics, 12 were not significantly expressed in any of the stages of steatosis (by adjusted p-value < 0.05) and were not included in visualization. We observed several genes with high effect size differences by steatosis grade, concordant with circulating proteomic and spatial relations (e.g., *IGFBP2*, *IL1RAP*, *SHBG*, *ENO3*, *DEGB1*, *ME1*). Across the genes prioritized by proteomic and spatial studies, we observed two types of discordant findings: (1) genes with a directionality consistent with our proteomic-spatial (but not bulk) results (e.g., *SERPINE1*/*PAI-1*^43–45^, *HSPA1B*^46^); (2) genes consistent with the bulk (but not proteomic-spatial) directionality (e.g., *PSAT1*^47^, *UDGH*^22^, *ACO1*^48^). Several factors—technical (sequencing methodologies, bulk versus single cell, limited sample size in this previously published scRNAseq dataset^34^, participant-level, and biological (steatosis as one component of the MASLD phenotype, in addition to inflammation, ballooning, fibrosis)—may account for these differences. Nevertheless, these findings collectively highlight the potential for proteo-transcriptional target mapping for human MASLD amidst context-dependent heterogeneity and complexity of integrating multiple approaches.

**Figure 6:**
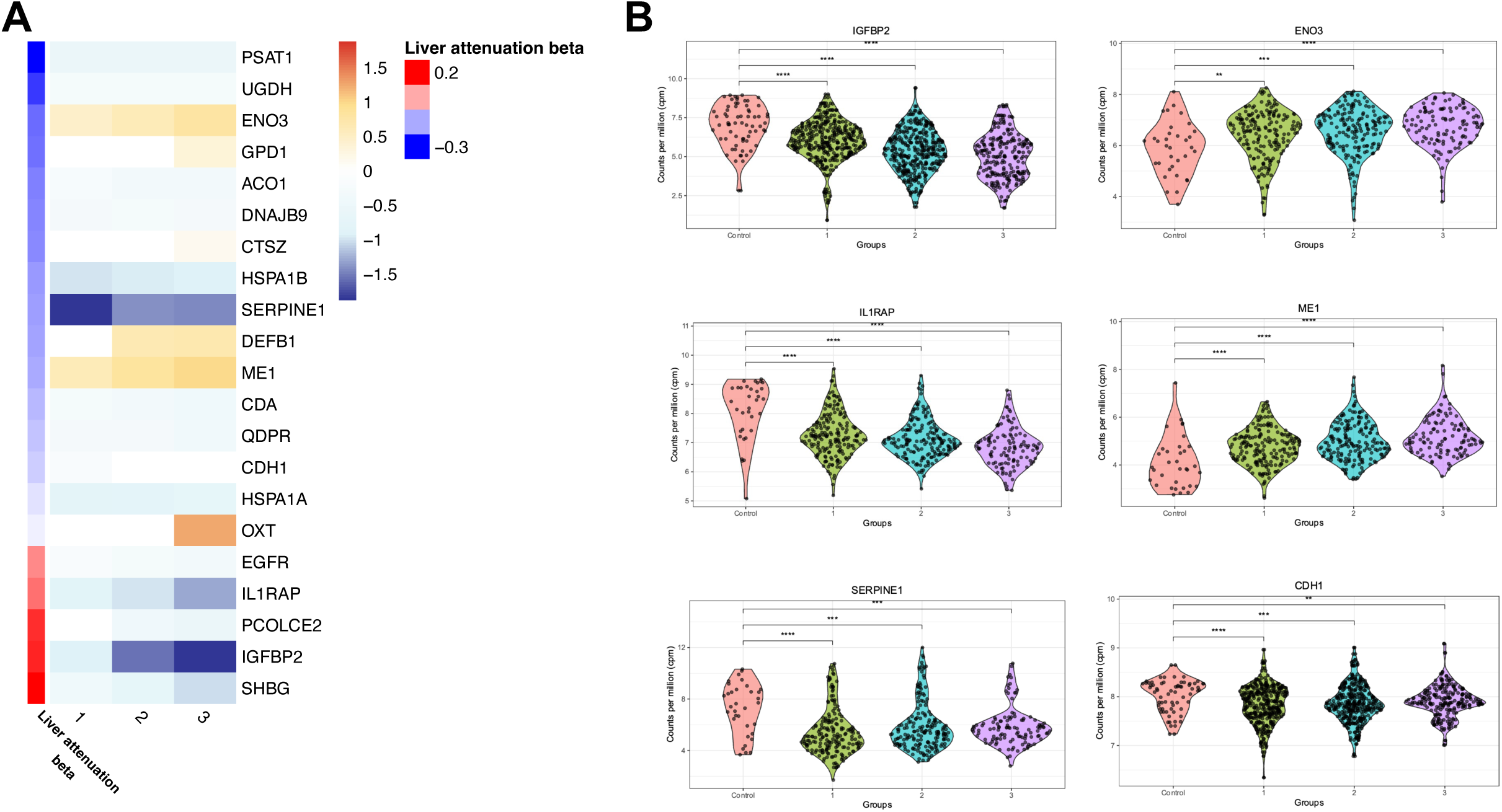
Transcriptional heterogeneity of spatial targets in human liver across steatosis stages. (**A**) Bulk transcript log2 fold change in human liver (over control samples without histologic steatosis) for those genes (among 33 significant on spatial studies) that were significantly differentially expressed in at least one comparison (stage 1 vs. control; stage 2 vs. control; stage 3 vs. control). Of the 33 genes passed forward for assessment in bulk transcriptomics, 12 were not significantly expressed in any of the stages of steatosis (by adjusted p-value < 0.05) and were not included in visualization. The “liver attenuation beta” represents the regression coefficient against liver attenuation in the CARDIA derivation sample. A positive coefficient (red) indicates a greater protein level is related to higher attenuation (lower steatosis); a negative coefficient (blue) indicates a greater protein level is related to lower attenuation (higher steatosis). This analysis excluded individuals with stage F4 fibrosis, given differences in hepatic physiology at this stage of decompensation. (**B**) Violin plots of example gene expression (in log2 counts per million) for genes that displayed a “concordant” directionality between the proteome and the bulk transcriptome (top and middle panel) and “discordant” directionality between proteome and bulk transcriptome.

### Humanized liver-on-a-chip recapitulates an early phase human MASLD phenotype consistent with proteo-transcriptional MASLD targets from human studies

The experimental design for the liver-on-a-chip (LOC) studies is shown in **Figures 7A-B**. Microscopy, immunofluorescence, and gene expression studies after administration of fatty acids (oleic and palmitic) known to promote a steatosis phenotype^49^ were consistent with morphologic and transcriptomic induction of a MASLD phenotype (decreased *IRS1*^50,51^, *IRS2*^51^, *PPARa*^52,53^; increased *SREBP1c*^54^, *PPARg*^55^, *FABP4*^56^; **Figure 7C-D**). Given limited cDNA yield from the LOC experiments, we prioritized 13 of 33 targets identified across proteomic and spatial transcriptional studies (**Supplemental Data File SD07**) for assessment on the LOC in two ways: (1) top 5 (*HMGCS1*, *SERPINE1*, *HSPA1B*, *ENO3*, *HSPA1A*) and bottom 5 (*CDA*, *PYGL*, *IL1RAP*, *SHBG*, *IGFBP2*) differentially expressed targets, ranked by log2-fold difference in steatotic and non-steatotic livers by spatial transcriptomics (include all 3 of which were downregulated in steatotic livers); (2) three additional targets differentially expressed in spatial human liver studies but with prominent expression in non-parenchymal (non-hepatocyte; NPCs) cells (*ME1*, *CTSZ*, *DEFB1*).

**Figure 7.**
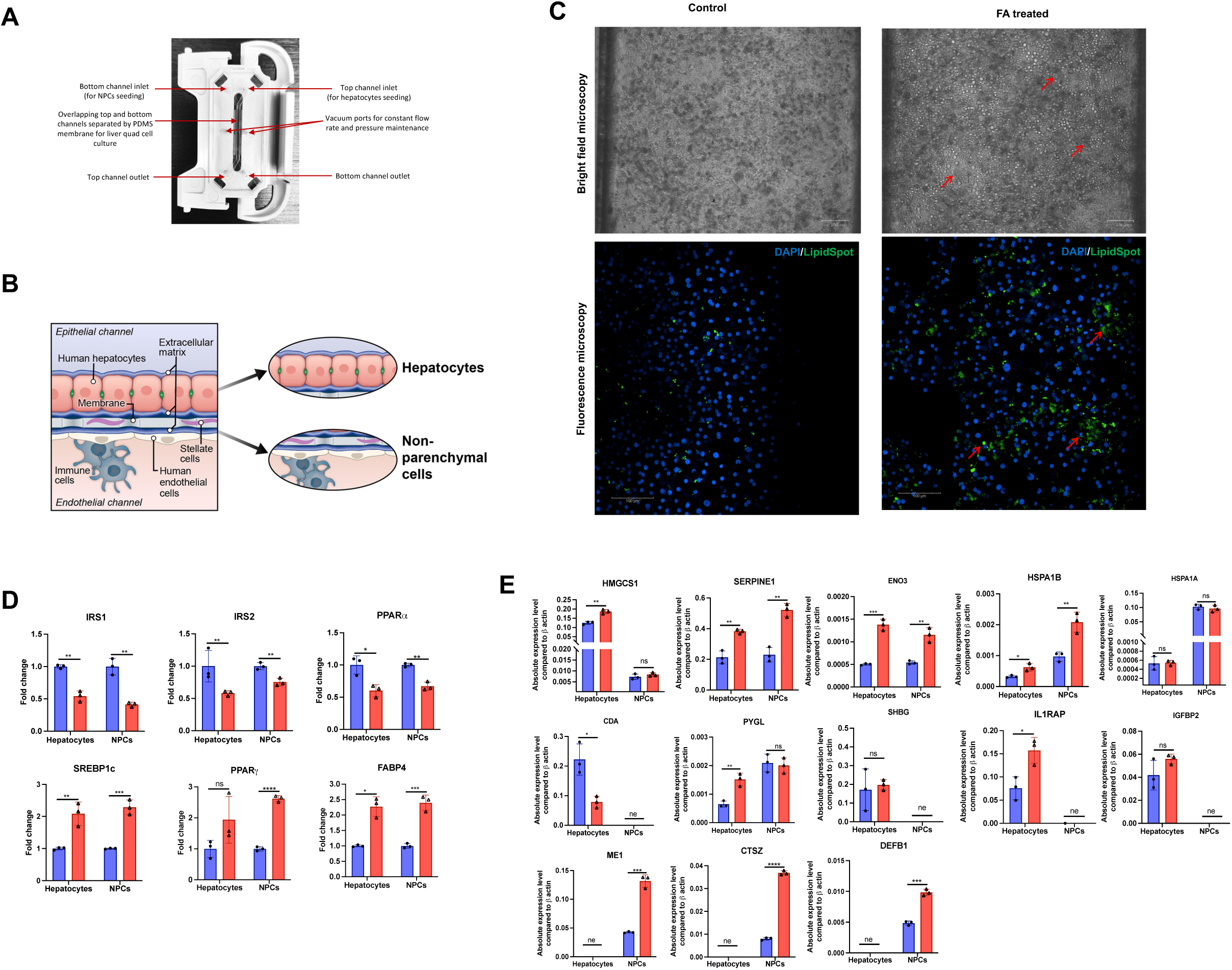
Transcriptional architecture of MASLD on a humanized liver-on-a-chip (LOC) largely replicates population and tissue findings. **(A)** and **(B)** show the structure and experimental design of MASLD induction on the LOC. **(C)** Successful MASLD model generation on a representative LOC. On the left, lipid droplet accumulation was visualized after 5-day treatment period of FAs. Representative brightfield and fluorescent confocal images of the LOC cells (scale bar = 100 μm). DAPI (nuclear) and LipidSpot (lipid droplet) stains are shown. Red arrows represent lipid droplet accumulation. **(D)** mRNA expression of canonical genes implicated in steatosis demonstrate expression patterns consistent with MASLD in both hepatocytes and NPCs. A total of 6 chips were included (3 FA and 3 control). Results were analyzed by an unpaired *t* test and expressed as mean ± standard error of 3 independent experiments. Each data point represents the average of 3 technical replicates. Control is in blue and FA treated is in red. Relative expression is shown as fold change (delta-delta CT) relative to control, normalized to beta-actin expression. **(E)** mRNA expression of top genes on the LOC that were prioritized by proteomic and transcriptomic studies (see text). Breaks in Y axis are presented given disparate expression of some genes (e.g., HMGCS1 had low expression in non-hepatocytes, while HSPA1A was expressed at low levels in hepatocytes). ME1, CTSZ and DEFB1 were not expressed in hepatocytes; CDA, SHBG, IL1RAP and IGFBP2 were not expressed in NPCs. See text for details. Abbreviations: *ne*, not expressed (raw Ct > 40); *ns*, non-significant; *, p<0.05; **, p<0.01; ***, p< 0.001; ****, p<0.0001.

We observed broadly directionally consistent results between circulating proteomic, tissue transcriptional, and LOC experiments, with increased expression of genes implicated in hepatic lipid metabolism, stress, and non-canonical pathways (e.g., ferroptosis) across hepatocytes and NPCs (*PYGL*, *HMGCS1*, *SERPINE1*, *ENO3*, *HSPA1B*; **Figure 7E**). Furthermore, while *ME1*, *CSTZ*, and *DEFB1* were not expressed in LOC hepatocytes (consistent with spatial human studies), the expression of these genes was increased in NPCs in the LOC (**Figure 7E**). Taken together with bulk data for *DEFB1* and *ME1* in bulk transcriptional data (**Figure 6**), these results suggest the increased expression of these genes are predominantly driven by cells of non-hepatocyte origin. Of note, several genes did not exhibit the expected directionality from proteomic, spatial, or bulk studies (non-significant: *IGFBP2*, *SHBG*; opposite directionality, *IL1RAP*, *CDA*; **Figure 7E**), potentially owing to biological heterogeneity between model systems.

## DISCUSSION

Given the growing worldwide prevalence of MASLD and its broad impact on human health, there is increasing interest to identify actionable biomarkers that reflect clinical risk and mechanism underlying MASLD pathobiology. We identified MASLD-associated proteins across a broad proteomic space (7230 aptamers) in ≈5000 individuals from two large prevention populations at dramatically lower metabolic risk relative to recent reports^6,8,57^ to address clinical generalizability. Using both single and multivariable regression modeling, the MASLD proteome prioritized broad metabolic, inflammatory, and fibrosis pathways, with multivariable instruments offering significant augmentation in MASLD discrimination beyond clinical factors in both CARDIA and UK Biobank (despite heterogeneity in the assessments of proteins, hepatic steatosis, and social-environmental measures). Our approach was strengthened by using direct measures of hepatic steatosis (CT, ultrasound, and MRI), as opposed to the use of diagnosis codes^58^, to define proteomic relations to MASLD, resulting in multivariable MASLD proteomic instruments associated with hepatic steatosis in populations with elevated metabolic risk (CCHC). The range in C-statistic for composite proteomic-phenotype risk models in both cohorts (CARDIA: 0.94; UK Biobank: 0.83) were consistent with prior reports in markedly higher risk individuals from Wood and colleagues (validation N=134 adults; BMI 49±8 kg/m^2^, 41% diabetes, 84% female; proteomic C-statistic: 0.864)^57^ and Govaere and colleagues (validation N=115 adults; BMI 32±6 kg/m^2^, 52% diabetes, 44% female; proteomic C-statistic: 0.8)^8^. The multivariable MASLD proteome forecasted prospective disease risk in >26000 UK Biobank participants for incident chronic non-alcoholic liver disease, diabetes, and other outcomes linked to MASLD (ischemic heart disease, cancer mortality, and all-cause mortality) and mitigated high polygenic liability for diabetes in UK Biobank. These results extend reports in high-risk, established disease populations^6^ into a prevention context with high clinical impact of intervention, underscoring the translatability of this approach for MASLD- and MASLD-related disease personalization.

While liver imaging enjoys discriminative accuracy for MASLD (e.g., CT, MRI)^59,60^, these studies have some limitations, including clinical availability and cost in widespread screening efforts and sensitivity for early disease where intervention may have the greatest clinical impact. Consequently, the clinical field in MASLD has directed focus toward identifying rapidly measured biosignatures of dynamic hepatic states that are prognostically, diagnostically, and biologically relevant. Candidate biomarkers have included human genetics^58,61–66^, gene and protein expression^6,58,67,68^, and metabolism. Nevertheless, while key genetic susceptibility loci critical to MASLD biology have been identified in large biobanks (e.g., *PNPLA3*), polygenic liability for MASLD in a population context is limited^69^ by more prominent non-genetic influences (e.g., diet, alcohol or drug exposure, obesity, inactivity, environmental exposures). In non-genomic studies across a broader array of biomarkers, sample sizes have generally been small with population bias (e.g., measured in established risk, like obesity^61^). Of note, recent innovative efforts to map a circulating snapshot of metabolic biology (via the human proteome) into hepatic transcriptional states has been successful, albeit in a small sample with high metabolic disease prevalence^8^. Indeed, the value of transcriptional indexing of the human proteome across broad at-risk populations has recently been highlighted^70^ (though not yet in MASLD).

Beyond disease personalization, our central hypothesis included mapping of the circulating human proteome into tissue MASLD to identify relevant, dynamic markers of interest. We observed a very high enrichment of transcript expression of genes encoding the MASLD proteome in the liver (far beyond any other tissue), supportive of its hepatic origin. Moreover, identified protein targets specified broad pathways central to MASLD, including regulation of hepatocyte regeneration (*EGFR*^23^), injury, apoptosis (*MET*^25^), inflammation (*CXCL2*^26^, *CRP*, *SERPINE1*), metabolism (*ACY1*^6,11^, *FAH*^12^, *ADH1A*^13,14^, *ALDOB, SORD*^15^, *AKR1D1*^16^, *AKR1C4*^17,18^), and fibrosis (*IGF-1*^24^). Given that these tissue references are from “normal” banks in bulk resolution (e.g., Human Protein Atlas/GTEX), we further explored our MASLD-associated proteomic targets at single nuclear and spatial resolution in human liver at an early MASLD stage^34^. A prime finding from this approach was the striking concordance of the <u>circulating</u> <u>proteomic</u> effect size in our large, at-risk population (CARDIA) and the fold-difference between healthy and fatty liver. Plasma proteins that were more abundant in patients with lower degree of hepatic steatosis corresponded to mRNAs that were also higher in expression in non-steatotic livers (and vice versa). Gene activity (as defined by a gene expression score across 198 MASLD-associated proteins) mapped primarily to histopathologic areas of steatosis, with a predominant hepatocyte expression and zonation pattern. Interestingly, we also observed a greater expression of *CTSZ* in macrophages, and migratory and conventional dendritic cells, which is in line with recent reports linking inflammatory cells to MASLD pathogenesis in a murine model^71^. Several genes differentially expressed in spatial transcription were dynamic across MASLD stages with consistent directionality with spatial transcriptomic and circulating proteomics ^7^, further suggesting validity (and prioritization) of these targets.

Our studies were augmented by the development of a MASLD model in a humanized LOC platform to determine direct causality between induction of steatosis and changes in mRNA transcripts identified in the human transcriptional studies. While the LOC model used in this study has been validated for recapitulating key aspects of human liver physiology^72–74^, it has mostly been used as a drug screen for hepatotoxicity^75–78^. While the use of such platforms to model steatosis is emerging^79^, we successfully developed a model that includes both hepatocytes and non-hepatocyte cells (Kupffer cells, stellate cells, and endothelial cells) subjected to treatment with a cocktail of fatty acids. Importantly, while admittedly far less complex than human MASLD, the model recapitulates key histological and transcriptional features of human MASLD, allowing us to query for direct changes (in response to steatosis induction) in mRNA transcripts in both hepatocyte and non-hepatocyte cell types derived from our human studies. These data not only validate a primary role for intrahepatic steatosis in the transcriptional changes, but also provide a possible platform to test novel therapeutic targets (that may reverse these changes) in the future.

Overall, the shared findings across biologically plausible pathways relevant to MASLD pathogenesis—and the association of these mediators in circulation with long-term outcomes— supports a broad validity from human populations to individual cells. Several caveats merit mention. We recognize the diagnosis of MASLD requires additional metabolic dysfunction (independent of other causes, e.g., viral, alcohol) associated with steatosis, and different cohorts had different distributions of these influences. Nevertheless, our CARDIA derivation necessarily excluded individuals with viral or drug/toxin-induced liver disease as possible in an epidemiologic setting and generated proteomic signatures associated with hepatic steatosis across BMI (a key determinant of metabolic dysfunction). Broad validation across cohorts (despite heterogeneity in hepatic steatosis ascertainment) is further evidence of external validity. The use of the transcriptome for tissue deconvolution of the human proteome is an emerging concept^8,80^, traditionally countered by concerns over protein-to-transcript concordance (including epigenetic effects that may disconnect them) and tissue ubiquity of the transcripts corresponding to the circulating proteome. Nevertheless, recent work in high metabolic risk individuals has suggested a largely positive (though variable) correlation between tissue RNA and circulating protein levels for those targets associated with MASLD stage^8^, consistent with our findings of a high liver transcriptional enrichment for prioritized proteins. Consistent cross-platform imaging and prognostic findings (e.g., aptamer- to antibody-based proteomics) lend validity. While the relation between the MASLD score and MRI based hepatic steatosis in UK Biobank were temporally separated, the association was similar as in CARDIA. We are limited in not having conducted spatial transcriptomics across variable disease states over time or with therapy (e.g., weight reduction), as well as gain- and loss-of-function studies for targets that survived our tiered approach. While this was not the scope of the current work, we envision that the approach here can be extended to a serial context to examine tissue-circulating concordance to further hone targets for mechanistic study. Finally, we noted some discordances between proteomic, spatial, bulk, and LOC data. Beyond technical considerations (e.g., bulk vs. spatial transcriptional approaches), we hypothesize variability in results may owe to population and disease heterogeneity across cohorts, including MASLD phenotypic heterogeneity (ballooning, inflammation, fibrosis, steatosis that can co-existent to different extent).

Nevertheless, targets that filter across all approaches are more likely to be involved in human liver disease, and larger, integrative approaches will continue to add to this science.

In conclusion, across β5000 participants with clinical, imaging, and biochemical data, we define a proteomic architecture of MASLD with replication and diagnostic stratification across a MASLD spectrum (from early- to high-risk metabolic cohorts) and strong association with MASLD-related disease (beyond modern human genetic approaches) in >26,000 individuals. Proteins implicated by these population-based approaches were highly enriched at a transcriptional level in human liver and specified canonical and novel pathways of MASLD progression. We observed spatially enriched activity of these genes in areas of steatosis and by liver zonation, with concordance between the circulating proteomic effects on liver fat and the fold differences between healthy and fatty liver by spatial transcription. Several targets additionally demonstrated concordant changes during evolution of MASLD across histologically defined stages and within a humanized “liver-on-a-chip” model system. These results contextualize the promise of multi-level discovery—across broad clinical populations, proteome, and tissue studies—to discern biologically relevant, spatially enriched targets in MASLD for downstream mechanistic, diagnostic, and prognostic work.

## METHODS

### Data Availability

Data for reproduction of this analysis may be obtained from the CARDIA Coordinating Center (www.cardia.dopm.uab.edu), CCHC Coordinating Center (https://sph.uth.edu/research/centers/hispanic-health/), SteatoSITE (https://steatosite.com), Liver Cell Atlas (https://livercellatlas.org/index.php), and UK Biobank (https://www.ukbiobank.ac.uk). Analyses in the UK Biobank were performed under proposal number 57492.

### Study samples

The study involved multiple samples: (1) the Coronary Artery Risk Development in Young Adults study (CARDIA, N=2679; proteomic discovery and validation of MASLD-related proteins; characteristics in **Supplementary Table 1**, study design in reference); (2) the UK Biobank study (N=26421; second validation of MASLD-related proteins; assessment of clinical prognostic value against incident MASLD-related diseases; characteristics in **Supplementary Table 2**); (3) Cameron County Hispanic Cohort (CCHC; N=206 with ultrasound-based measures of liver structure and circulating proteomics; characteristics in **Supplementary Table 3**) (4) published bulk RNA sequencing study (SteatoSITE, N=618 liver biopsies across stages of MASLD)^7^; (5) spatial transcriptomic study in human liver (N=5 liver biopsies for spatial transcriptomics and N=19 for single nuclear RNA-sequencing)^34^. All study participants provided written and informed consent, and all study protocols were approved by the Institutional Review Boards of the respective studies.

#### CARDIA

The Coronary Artery Risk Development in Young Adults (CARDIA) study started recruitment in 1985-1986 across 4 cities in the U.S. (Birmingham, AL; Chicago, IL; Minneapolis, MN; and Oakland, CA) to study coronary risk factor development longitudinally beginning in young adulthood^81–84^. Our study used data from the Year 25 exam where 2977 participants had proteomics quantified. We excluded 275 participants with other potential causes for hepatics steatosis (>14 alcoholic drinks/week, hepatitis C, cirrhosis, HIV, and use of amiodarone, valproic acid, methotrexate, tamoxifen, or diltiazem)^10^. We excluded 11 participants missing hepatic steatosis measurements, and 12 participants for missing data on BMI or drinks/week. CARDIA participants were randomly split into derivation (70%) and validation (30%) samples, balanced by computed tomography-based measurement of hepatic steatosis.

#### UK Biobank

The UK Biobank is a population-based study of >500000 participants who were aged 40-69 when recruited between 2006-2010 across the United Kingdom. Proteomics data from the initial assessment (instance 0) using the Olink Explore panel is available on β54000 UK Biobank participants^80^. We included 26429 participants with complete data for the proteins used to calculate a proteomic score of hepatic steatosis, of which 8 participants were excluded from analyses for having a proteomic score >5 SDs away from the mean. A subset of 2111 had hepatic steatosis quantified by MRI at the imaging visit (2014 and later; instance 2)

#### Cameron County Hispanic Cohort (CCHC)

The CCHC is a community-based prospective observational cohort study of 5122 individuals (age 8-90) from a low-income Hispanic/Latino population at the Texas/Mexico border. The study design has been previously described^85^. We included 206 individuals who had abdominal ultrasound to measure controlled attenuation parameter (CAP), a quantitative measure of hepatic steatosis^86^, and simultaneous circulating proteomics.

### Hepatic steatosis assessment

In CARDIA, hepatic steatosis was measured as liver attenuation on computed tomography as previously described, where lower levels of liver attenuation are associated with greater steatosis^10^. MASLD was defined as liver attenuation <40 Hounsfield units. In the UK Biobank, hepatic steatosis was measured by magnetic resonance imaging in a subset of participants at instance 2 using the iterative decomposition of water and fat with echo asymmetric and least-squares estimation (IDEAL) protocol, as previously described^87^. MASLD was defined as a proton density fat fraction >5.5%^32^. In CCHC, vibration-controlled transient elastography was used to measure CAP (FibroScan 502 Touch or FibroScan 530 Compact, Echosens; automatic probe selection) for 10 valid measures with the median used in analysis, as described^88^.

### Proteomics

#### CARDIA

Quantification of the circulating proteome was performed using aptamer-based technology (Somalogic, Boulder, CO) which measured 7596 aptamers. Sixty-eight participants had >1 samples measured and we averaged their proteomic data for analysis. We excluded non-human proteins (N=72) and proteins with a coefficient of variation >20% (N=58). We tested for batch effect and participant outliers using principal component analysis and identified neither. Proteins were log-transformed and standardized (mean 0, variance 1) prior to use in models.

#### UK Biobank

Recently released proteomic data from the Olink Explore platform (Olink, Uppsala, Sweden) measured from the instance 0 visit were used in this study^80^. Of the 1463 proteins measured, we excluded 130 proteins where >40% of reported measurements were below the limit of detection and another 3 proteins where >20% of reported measurements were missing. Proteins were standardized (mean 0, variance 1) prior to use in models.

#### CCHC

We performed proteomics in CCHC participants using the Olink Explore 1536 platform. Proteins were standardized (mean 0, variance 1) prior to use in models.

### Spatial, single nuclear, and bulk transcriptomics in human liver

#### Single nuclear and spatial transcriptomics

To assess cell-specific and spatial expression patterns of implicated protein targets, we harnessed integrated single cell and single nuclear RNA-sequencing (scRNA seq, total N= 19; fatty = 7; non-fatty = 11; unknown = 1) and Visium spatial transcriptomics data (total N= 4; fatty = 2; non-fatty = 2) previously published from our collaborative group^34^. Expression patterns of implicated proteins were assessed by mapping significant model proteins to their corresponding gene symbol that were expressed in both the scRNA-seq and Visium data, resulting in total of 198 genes represented across the three datasets. Activity of these genes were then measured for their activity using single gene expression measures as wells an expression composite score that represent the transcriptional signature of all model genes in each individual cell (snRNAseq) or spot (Visium data).

Expression composite score was generated using the *AddModuleScore* function (implemented in Seurat v5). To identify differential expression of nominated targets in the liver we compared healthy samples to early steatotic samples using Visium data where both healthy and early steatotic samples were available. Differentially expressed genes were assessed using negative binomial model implemented in the *FindMarkers* function (Seurat). Only target genes expressed in at least 10% of the spots were included in the analysis (198 genes) for differential expression. We defined differential expression as adjusted p-value < 0.05 and |log2fold change > 0.25| and a minimum difference in expressed spots > 10% between fatty and non-fatty. We confirmed the effect size estimates of our differential expression analysis via negative binomial mixed models with sample as random effect (generalized linear mixed models are more sensitive to the dispersion in single-cell data compared to generalized linear models), with high agreement between log2 fold change and the negative binomial mixed model coefficients for all 198 model targets (Pearson r =0.86) and for the 33 differentially expressed genes (Pearson r =0.99; **Supplemental Figure 6**).

#### Transcriptional differences across MASLD stages

We explored the pattern of expression across the 33 genes prioritized by the spatial data analysis above (33 differentially expressed genes between healthy and steatotic tissue out of 198 genes tested, see **Results**) in the SteatoSITE cohort (523 samples; 34 controls, 489 with MASLD) categorized based on NAFLD activity score (NAS) for steatosis (only those samples with scores 1, 2 and 3 were chosen) and compared to control samples^7^. We excluded those individuals with NASH-CRN stage F4 fibrosis, given differences in expression patterns detected in our initial study^7^ and differences in physiology with advanced fibrosis (including paradoxical loss of hepatic fat^89^). Reads were normalised using the weighted trimmed mean of M values method^90^. Differential gene expression analysis was performed using *limma-voom* (v3.28.14) with the protein-coding genes using an FDR of 5% (Benjamini-Hochberg)^91^. Of the 33 genes passed forward for assessment in bulk transcriptomics, 12 were not significantly expressed in any of the stages of steatosis (by adjusted p-value < 0.05) and were not included in visualization.

#### Humanized liver-on-a-chip MASLD model

The goal of “liver-on-a-chip” technology is to simulate the liver microenvironment which retains key characteristics of native liver function over long-term in culture. The quad culture was set up following the manufacturer’s protocol.

The methods below are reproduced from our recent work^92^ directly for rigor and reproducibility, and this citation provides scientific attribution for this. Briefly, by design, each polydimethylsiloxane (PDMS) chip (Chip-S1; Emulate) includes hepatocytes in the apical channel and non-parenchymal cells [NPCs: Kupffer, Stellate and Liver Sinusoidal Endothelial Cells (LSECs)] in the basal channel (**Supplementary Table 4**). These two channels are separated by a porous membrane, coated by hepatic extracellular matrix (ECM). This setting allows the cell-to-cell interaction mimicking the *in vivo* system. The top channel was seeded with hepatocytes at a concentration of 3.5×10^6^ cells/mL, followed by overlay with matrigel on the next day. The day after hepatocyte overlay, cell suspensions of three NPCs were mixed in a 1:1:1 ratio (v/v/v) to generate the bottom channel tri-cell mixture. The final seeding density of each cell types in the bottom channel were: LSECs: 3×10^6^ cells/mL; Stellate cells: 0.1×10^6^ cells/mL; Kupffer cells: 0.5×10^6^ cells/mL. Chips were maintained for another 96 hours at this condition before treating with fatty acids (FAs). We mimicked an early phase of MASLD by treating both channels of the LOC either with vehicle control (1% BSA) or a combination of FAs (oleic acid 300μM: 300μM palmitic acid bound with 1% BSA) for 5 consecutive days.

Hepatocytes and NPCs treated with either vehicle control or FAs were imaged directly under brightfield microscope (BioRad). Chips were fixed with 4% paraformaldehyde (4%PFA) followed by permeabilization of both channels with 0.1% Triton® X-100 before staining. Permeabilized cells in both channels were incubated with LipidSpot™ for 10 minutes. The chips were examined under a fluorescence microscope (ECHO Revolve microscope).

After 5 days of dosing with FAs, the chips were disconnected, washed with 1X PBS, and filled with RNAlater (Invitrogen) to preserve cells for RNA extraction. The PureLink RNA Mini Kit (Thermo Fischer Scientific) was used following the manufacturer’s protocol. Total RNA was eluted in 20µL, treated with DNAse, and “cleaned-up” using RNA Clean & Concentrator-5 with DNase I (Zymo Research) following manufacturer’s protocol. Final RNA concentration was quantified by spectrophotometry (Nanodrop 2000, Thermo Fischer Scientific). The High-Capacity cDNA Reverse Transcription Kit (Thermo Fischer Scientific) was used for cDNA synthesis from RNA. For amplification and quantification of selected genes (*HMGCS1*, *SERPINE1*, *HSPA1B*, *ENO3*, *HSPA1A*, *PYGL*, *CDA*, *SHBG*, *IL1RAP*, *IGFBP2*, *ME1*, *CTSZ*, *DEFB1*, *IRS1*, *IRS2*, *FABP4*, *SREBP1c*, *PPARα*, *PPARγ*, and β-actin) the ExiLENT SYBR® Green master mix (Exiqon, Vedbæk) was used on a Quant Studio 6 Flex Real-Time PCR System up to 40 amplification cycles. Any amplification cycle (Ct) greater than or equal to 40 was assigned as a “negative threshold”, which means the corresponding genes were not expressed above the limit of detection of the qRT-PCR assay and therefore those genes were not included in our calculations. For **Fig 7E** absolute gene expression was quantified by 2^-ΔCt^ method after normalization of genes of interest to the internal control β-ACTIN, whereas relative gene expression was used for **Fig 7D**. All qRT-PCR primer sequences are summarized in **Supplementary Table 5**.

### Statistical methods

#### Relating the circulating proteome to hepatic steatosis to identify biological pathways of steatosis and development of a diagnostic biomarker panel

Relations of individual aptamers with hepatic steatosis were examined via regression with aptamers as the predictors adjusted for age, sex, race, and BMI with a false discovery rate of 5% (Benjamini-Hochberg) in the CARDIA cohort using a derivation (70%) and validation (30%) split design balanced on CT liver attenutation. To generate a multivariable protein score of hepatic steatosis (referred to as “MASLD score”), we used least absolute shrinkage and selection operator (LASSO) with non-penalized adjustments for age, sex, race, and BMI in the CARDIA derivation sample, and replicated its relation in the CARDIA validation sample. This MASLD score was then recalibrated for use in UK Biobank and CCHC (which used Olink proteomics platforms in contrast to CARDIA, which used a SomaScan platform), using LASSO regression with the original MASLD score as the dependent variable and all overlapping proteins (matching between the Olink and SomaScan platforms on UniProt identifier) as the independent variables. Pathway analysis was performed on proteins that were significant in both CARDIA derivation and validation subsamples (FDR<5%) using R package ClusterProfiler^93^ on KEGG and Reactome database. Hypergeometric tests were used to evaluate enrichment level for each pathway utilizing all proteins on the SomaScan platform as background. The top 10 most enriched pathways in both KEGG and Reactome were visualized together via lollipop plots. To identify hub genes, Protein-protein interactions for 235 significant genes were retrieved from the STRING database^94^. Hub genes were determined as any protein with more than 5 high-confidence interactions (score>700) and hub genes and all interactions were visualized using Cytoscape^95^. Tissue-specific gene expression enrichment was performed by R package TissueEnrich^96^ based on tissue expression patterns in Human Protein Atlas database^97^. To examine the clinical utility of the MASLD score as a diagnostic marker of MASLD, we compared a clinical model of MASLD (age, sex, race, BMI, alcoholic drinks per week, AST, ALT, hemoglobin A1c) to a model with the MASLD score using receiver operator curve area under the curve analysis in both the CARDIA validation sample and UK Biobank.

#### Testing the association between a protein score of MASLD with development of MASLD and clinical outcomes

In UK Biobank, Cox regression was used to examine the relation of the MASLD score with clinical endpoints. Death and type of death (cardiovascular death, cancer death, respiratory death) were defined by using death registry data (UK Biobank Data Field 40000) in conjunction with the primary cause of death International Classification of Disease (ICD) 10 code (UK Biobank Data Field 40001). Translating ICD10 codes to type of death was conducted as previously reported^98^. Censoring for clinical endpoints was determined by region-specific censor dates for each participant based on the location of initial assessment (UK Biobank Data Field 54). Deaths were censored on 30 November 2022 for all participants. Non-death outcomes in UK Biobank were defined by ICD10 diagnosis codes grouped into relevant “phecodes” via the PheWAS package^99^. For each phecode, we generated a case, control, and excluded status for each subject. Time to event for phecodes was defined as the date of the earliest relevant ICD10 was documented. Prevalent conditions were defined by self-report or physician diagnosis (Data Fields 20002, 2443, 6150). Sequential models with increasing adjustments were created 1) unadjusted 2) age, sex, race, BMI 3) age, sex, race, BMI, Townsend Deprivation Index, diabetes, smoking, alcohol use, systolic blood pressure, and LDL. We conducted a sensitivity analysis including further adjustment for AST, ALT, and hemoglobin A1c. We compared adjusted models (age, sex, race, BMI, diabetes [removed from models for diabetes], smoking, alcohol use, systolic blood pressure, LDL) with and without the MASLD score to compare differences in C-statistics and net reclassification index (NRI; calculated at the 75th percentile for NRI for events). To investigate the effect of the MASLD score on polygenic risk, we examined the relationship between the MASLD score with the standard polygenic risk score (PRS) of diabetes (Data Field 26285) in a Cox model for incident type 2 diabetes as a function of the MASLD score and the PRS for type 2 diabetes, including an interaction term, with adjustments for age, sex, race, and the top 4 principal components of genetic ancestry.

## ACKNOWLEDGEMENTS

CARDIA is conducted and supported by the NHLBI in collaboration with the University of Alabama at Birmingham (HHSN268201800005I & HHSN268201800007I), Northwestern University (HHSN268201800003I), University of Minnesota (HHSN268201800006I), and Kaiser Foundation Research Institute (HHSN268201800004I). Proteomics quantification was funded by the NHLBI (HL122477; PI Kalhan). This manuscript has been reviewed by CARDIA for scientific content. The views expressed in this manuscript are those of the authors and do not necessarily represent the views of the NHLBI; the NIH; or the U.S. Department of Health and Human Services. The authors would like to thank the CCHC cohort team, particularly Rocío Uribe who recruited and interviewed the participants. Marcela Morris, BS, and Hugo Soriano and their teams for laboratory and data support respectively; Norma Pérez-Olazarán, BBA, and Christina Villarreal, BA for administrative support; Valley Baptist Medical Center, Brownsville, Texas, for providing us space for our Center for Clinical and Translational Science Clinical Research Unit is located; and the community of Brownsville and the participants who so willingly participated in this study in their city. This study was funded in part by Center for Clinical and Translational Sciences, National Institutes of Health Clinical and Translational Award grant no. UL1 TR000371 from the National Center for Advancing Translational Sciences.

## REFERENCES

1. Riazi K, Azhari H, Charette JH, Underwood FE, King JA, Afshar EE, Swain MG, Congly SE, Kaplan GG, Shaheen AA. The prevalence and incidence of NAFLD worldwide: a systematic review and meta-analysis. Lancet Gastroenterol Hepatol. 2022;7:851–861. doi: 10.1016/S2468-1253(22)00165-0

2. Anstee QM, Reeves HL, Kotsiliti E, Govaere O, Heikenwalder M. From NASH to HCC: current concepts and future challenges. Nat Rev Gastroenterol Hepatol. 2019;16:411–428. doi: 10.1038/s41575-019-0145-7

3. Romeo S, Kozlitina J, Xing C, Pertsemlidis A, Cox D, Pennacchio LA, Boerwinkle E, Cohen JC, Hobbs HH. Genetic variation in PNPLA3 confers susceptibility to nonalcoholic fatty liver disease. Nature genetics. 2008;40:1461–1465. doi: 10.1038/ng.257

4. Luo Y, Wadhawan S, Greenfield A, Decato BE, Oseini AM, Collen R, Shevell DE, Thompson J, Jarai G, Charles ED, Sanyal AJ. SOMAscan Proteomics Identifies Serum Biomarkers Associated With Liver Fibrosis in Patients With NASH. Hepatol Commun. 2021;5:760–773. doi: 10.1002/hep4.1670

5. Corey KE, Pitts R, Lai M, Loureiro J, Masia R, Osganian SA, Gustafson JL, Hutter MM, Gee DW, Meireles OR, et al. ADAMTSL2 protein and a soluble biomarker signature identify at-risk non-alcoholic steatohepatitis and fibrosis in adults with NAFLD. Journal of hepatology. 2022;76:25–33. doi: 10.1016/j.jhep.2021.09.026

6. Sanyal AJ, Williams SA, Lavine JE, Neuschwander-Tetri BA, Alexander L, Ostroff R, Biegel H, Kowdley KV, Chalasani N, Dasarathy S, et al. Defining the serum proteomic signature of hepatic steatosis, inflammation, ballooning and fibrosis in non-alcoholic fatty liver disease. Journal of hepatology. 2023;78:693–703. doi: 10.1016/j.jhep.2022.11.029

7. Kendall TJ, Jimenez-Ramos M, Turner F, Ramachandran P, Minnier J, McColgan MD, Alam M, Ellis H, Dunbar DR, Kohnen G, et al. An integrated gene-to-outcome multimodal database for metabolic dysfunction-associated steatotic liver disease. Nat Med. 2023;29:2939–2953. doi: 10.1038/s41591-023-02602-2

8. Govaere O, Hasoon M, Alexander L, Cockell S, Tiniakos D, Ekstedt M, Schattenberg JM, Boursier J, Bugianesi E, Ratziu V, et al. A proteo-transcriptomic map of non-alcoholic fatty liver disease signatures. Nat Metab. 2023;5:572–578. doi: 10.1038/s42255-023-00775-1

9. Niu L, Thiele M, Geyer PE, Rasmussen DN, Webel HE, Santos A, Gupta R, Meier F, Strauss M, Kjaergaard M, et al. Noninvasive proteomic biomarkers for alcohol-related liver disease. Nature medicine. 2022;28:1277–1287. doi: 10.1038/s41591-022-01850-y

10. VanWagner LB, Wilcox JE, Ning H, Lewis CE, Carr JJ, Rinella ME, Shah SJ, Lima JAC, Lloyd-Jones DM. Longitudinal Association of Non-Alcoholic Fatty Liver Disease With Changes in Myocardial Structure and Function: The CARDIA Study. J Am Heart Assoc. 2020;9:e014279. doi: 10.1161/JAHA.119.014279

11. Pirola CJ, Sookoian S. Multiomics biomarkers for the prediction of nonalcoholic fatty liver disease severity. World journal of gastroenterology: WJG. 2018;24:1601–1615. doi: 10.3748/wjg.v24.i15.1601

12. Ma J, Tan X, Kwon Y, Delgado ER, Zarnegar A, DeFrances MC, Duncan AW, Zarnegar R. A Novel Humanized Model of NASH and Its Treatment With META4, A Potent Agonist of MET. Cell Mol Gastroenterol Hepatol. 2022;13:565–582. doi: 10.1016/j.jcmgh.2021.10.007

13. Li H, Toth E, Cherrington NJ. Alcohol Metabolism in the Progression of Human Nonalcoholic Steatohepatitis. Toxicol Sci. 2018;164:428–438. doi: 10.1093/toxsci/kfy106

14. Aljomah G, Baker SS, Liu W, Kozielski R, Oluwole J, Lupu B, Baker RD, Zhu L. Induction of CYP2E1 in non-alcoholic fatty liver diseases. Exp Mol Pathol. 2015;99:677–681. doi: 10.1016/j.yexmp.2015.11.008

15. Niu L, Geyer PE, Wewer Albrechtsen NJ, Gluud LL, Santos A, Doll S, Treit PV, Holst JJ, Knop FK, Vilsboll T, et al. Plasma proteome profiling discovers novel proteins associated with non-alcoholic fatty liver disease. Mol Syst Biol. 2019;15:e8793. doi: 10.15252/msb.20188793

16. Gathercole LL, Nikolaou N, Harris SE, Arvaniti A, Poolman TM, Hazlehurst JM, Kratschmar DV, Todorcevic M, Moolla A, Dempster N, et al. AKR1D1 knockout mice develop a sex-dependent metabolic phenotype. J Endocrinol. 2022;253:97–113. doi: 10.1530/JOE-21-0280

17. Zeng CM, Chang LL, Ying MD, Cao J, He QJ, Zhu H, Yang B. Aldo-Keto Reductase AKR1C1-AKR1C4: Functions, Regulation, and Intervention for Anti-cancer Therapy. Front Pharmacol. 2017;8:119. doi: 10.3389/fphar.2017.00119

18. Lyall MJ, Cartier J, Thomson JP, Cameron K, Meseguer-Ripolles J, O’Duibhir E, Szkolnicka D, Villarin BL, Wang Y, Blanco GR, et al. Modelling non-alcoholic fatty liver disease in human hepatocyte-like cells. Philos Trans R Soc Lond B Biol Sci. 2018;373. doi: 10.1098/rstb.2017.0362

19. Gorce M, Lebigot E, Arion A, Brassier A, Cano A, De Lonlay P, Feillet F, Gay C, Labarthe F, Nassogne MC, et al. Fructose-1,6-bisphosphatase deficiency causes fatty liver disease and requires long-term hepatic follow-up. J Inherit Metab Dis. 2022;45:215–222. doi: 10.1002/jimd.12452

20. Coles BF, Kadlubar FF. Human alpha class glutathione S-transferases: genetic polymorphism, expression, and susceptibility to disease. Methods Enzymol. 2005;401:9–42. doi: 10.1016/S0076-6879(05)01002-5

21. Nagamani SC, Erez A, Lee B. Argininosuccinate lyase deficiency. Genet Med. 2012;14:501–507. doi: 10.1038/gim.2011.1

22. Zhang T, Zhang N, Xing J, Zhang S, Chen Y, Xu D, Gu J. UDP-glucuronate metabolism controls RIPK1-driven liver damage in nonalcoholic steatohepatitis. Nat Commun. 2023;14:2715. doi: 10.1038/s41467-023-38371-2

23. Bhushan B, Banerjee S, Paranjpe S, Koral K, Mars WM, Stoops JW, Orr A, Bowen WC, Locker J, Michalopoulos GK. Pharmacologic Inhibition of Epidermal Growth Factor Receptor Suppresses Nonalcoholic Fatty Liver Disease in a Murine Fast-Food Diet Model. Hepatology. 2019;70:1546–1563. doi: 10.1002/hep.30696

24. Ohkubo R, Mu WC, Wang CL, Song Z, Barthez M, Wang Y, Mitchener N, Abdullayev R, Lee YR, Ma Y, et al. The hepatic integrated stress response suppresses the somatotroph axis to control liver damage in nonalcoholic fatty liver disease. Cell reports. 2022;41:111803. doi: 10.1016/j.celrep.2022.111803

25. Kroy DC, Schumacher F, Ramadori P, Hatting M, Bergheim I, Gassler N, Boekschoten MV, Muller M, Streetz KL, Trautwein C. Hepatocyte specific deletion of c-Met leads to the development of severe non-alcoholic steatohepatitis in mice. Journal of hepatology. 2014;61:883–890. doi: 10.1016/j.jhep.2014.05.019

26. Xiong X, Kuang H, Ansari S, Liu T, Gong J, Wang S, Zhao XY, Ji Y, Li C, Guo L, et al. Landscape of Intercellular Crosstalk in Healthy and NASH Liver Revealed by Single-Cell Secretome Gene Analysis. Molecular cell. 2019;75:644–660 e645. doi: 10.1016/j.molcel.2019.07.028

27. Del Ben M, Overi D, Polimeni L, Carpino G, Labbadia G, Baratta F, Pastori D, Noce V, Gaudio E, Angelico F, Mancone C. Overexpression of the Vitronectin V10 Subunit in Patients with Nonalcoholic Steatohepatitis: Implications for Noninvasive Diagnosis of NASH. International journal of molecular sciences. 2018;19. doi: 10.3390/ijms19020603

28. Schwabe RF, Tabas I, Pajvani UB. Mechanisms of Fibrosis Development in Nonalcoholic Steatohepatitis. Gastroenterology. 2020;158:1913–1928. doi: 10.1053/j.gastro.2019.11.311

29. Yang L, Sun Z, Li J, Pan X, Wen J, Yang J, Wang Q, Chen P. Genetic Variants of Glycogen Metabolism Genes Were Associated With Liver PDFF Without Increasing NAFLD Risk. Frontiers in genetics. 2022;13:830445. doi: 10.3389/fgene.2022.830445

30. Chella Krishnan K, Floyd RR, Sabir S, Jayasekera DW, Leon-Mimila PV, Jones AE, Cortez AA, Shravah V, Peterfy M, Stiles L, et al. Liver Pyruvate Kinase Promotes NAFLD/NASH in Both Mice and Humans in a Sex-Specific Manner. Cell Mol Gastroenterol Hepatol. 2021;11:389–406. doi: 10.1016/j.jcmgh.2020.09.004

31. Saed CT, Tabatabaei Dakhili SA, Ussher JR. Pyruvate Dehydrogenase as a Therapeutic Target for Nonalcoholic Fatty Liver Disease. ACS Pharmacol Transl Sci. 2021;4:582–588. doi: 10.1021/acsptsci.0c00208

32. Browning JD, Szczepaniak LS, Dobbins R, Nuremberg P, Horton JD, Cohen JC, Grundy SM, Hobbs HH. Prevalence of hepatic steatosis in an urban population in the United States: impact of ethnicity. Hepatology. 2004;40:1387–1395. doi: 10.1002/hep.20466

33. Khera AV, Chaffin M, Aragam KG, Haas ME, Roselli C, Choi SH, Natarajan P, Lander ES, Lubitz SA, Ellinor PT, Kathiresan S. Genome-wide polygenic scores for common diseases identify individuals with risk equivalent to monogenic mutations. Nat Genet. 2018;50:1219–1224. doi: 10.1038/s41588-018-0183-z

34. Guilliams M, Bonnardel J, Haest B, Vanderborght B, Wagner C, Remmerie A, Bujko A, Martens L, Thone T, Browaeys R, et al. Spatial proteogenomics reveals distinct and evolutionarily conserved hepatic macrophage niches. Cell. 2022;185:379–396 e338. doi: 10.1016/j.cell.2021.12.018

35. Faramia J, Hao Z, Mumphrey MB, Townsend RL, Miard S, Carreau AM, Nadeau M, Frisch F, Baraboi ED, Grenier-Larouche T, et al. IGFBP-2 partly mediates the early metabolic improvements caused by bariatric surgery. Cell Rep Med. 2021;2:100248. doi: 10.1016/j.xcrm.2021.100248

36. Lin YH, Wei Y, Zeng Q, Wang Y, Pagani CA, Li L, Zhu M, Wang Z, Hsieh MH, Corbitt N, et al. IGFBP2 expressing midlobular hepatocytes preferentially contribute to liver homeostasis and regeneration. Cell Stem Cell. 2023;30:665–676 e664. doi: 10.1016/j.stem.2023.04.007

37. Hedbacker K, Birsoy K, Wysocki RW, Asilmaz E, Ahima RS, Farooqi IS, Friedman JM. Antidiabetic effects of IGFBP2, a leptin-regulated gene. Cell metabolism. 2010;11:11–22. doi: 10.1016/j.cmet.2009.11.007

38. Chen X, Tang Y, Chen S, Ling W, Wang Q. IGFBP-2 as a biomarker in NAFLD improves hepatic steatosis: an integrated bioinformatics and experimental study. Endocrine connections. 2021;10:1315–1325. doi: 10.1530/EC-21-0353

39. Lu D, Xia Q, Yang Z, Gao S, Sun S, Luo X, Li Z, Zhang X, Han S, Li X, Cao M. ENO3 promoted the progression of NASH by negatively regulating ferroptosis via elevation of GPX4 expression and lipid accumulation. Annals of translational medicine. 2021;9:661. doi: 10.21037/atm-21-471

40. Wang J, Chen L, Li Y, Guan XY. Overexpression of cathepsin Z contributes to tumor metastasis by inducing epithelial-mesenchymal transition in hepatocellular carcinoma. PLoS One. 2011;6:e24967. doi: 10.1371/journal.pone.0024967

41. Zhang J, Fan N, Peng Y. Heat shock protein 70 promotes lipogenesis in HepG2 cells. Lipids in health and disease. 2018;17:73. doi: 10.1186/s12944-018-0722-8

42. Chen D, Dong X, Chen D, Lin J, Lu T, Shen J, Ye H. Cdh1 plays a protective role in nonalcoholic fatty liver disease by regulating PPAR/PGC-1alpha signaling pathway. Biochemical and biophysical research communications. 2023;681:13–19. doi: 10.1016/j.bbrc.2023.09.038

43. Henkel AS, Khan SS, Olivares S, Miyata T, Vaughan DE. Inhibition of Plasminogen Activator Inhibitor 1 Attenuates Hepatic Steatosis but Does Not Prevent Progressive Nonalcoholic Steatohepatitis in Mice. Hepatol Commun. 2018;2:1479–1492. doi: 10.1002/hep4.1259

44. Lee SM, Dorotea D, Jung I, Nakabayashi T, Miyata T, Ha H. TM5441, a plasminogen activator inhibitor-1 inhibitor, protects against high fat diet-induced non-alcoholic fatty liver disease. Oncotarget. 2017;8:89746–89760. doi: 10.18632/oncotarget.21120

45. Day K, Seale LA, Graham RM, Cardoso BR. Selenotranscriptome Network in Non-alcoholic Fatty Liver Disease. Frontiers in nutrition. 2021;8:744825. doi: 10.3389/fnut.2021.744825

46. Wang L, Zhou K, Wu Q, Zhu L, Hu Y, Yang X, Li D. Microanatomy of the metabolic associated fatty liver disease (MAFLD) by single-cell transcriptomics. J Drug Target. 2023;31:421–432. doi: 10.1080/1061186X.2023.2185626

47. Sim WC, Lee W, Sim H, Lee KY, Jung SH, Choi YJ, Kim HY, Kang KW, Lee JY, Choi YJ, et al. Downregulation of PHGDH expression and hepatic serine level contribute to the development of fatty liver disease. Metabolism. 2020;102:154000. doi: 10.1016/j.metabol.2019.154000

48. Kuwashiro S, Terai S, Oishi T, Fujisawa K, Matsumoto T, Nishina H, Sakaida I. Telmisartan improves nonalcoholic steatohepatitis in medaka (Oryzias latipes) by reducing macrophage infiltration and fat accumulation. Cell and tissue research. 2011;344:125–134. doi: 10.1007/s00441-011-1132-7

49. Moravcova A, Cervinkova Z, Kucera O, Mezera V, Rychtrmoc D, Lotkova H. The effect of oleic and palmitic acid on induction of steatosis and cytotoxicity on rat hepatocytes in primary culture. Physiol Res. 2015;64:S627–636. doi: 10.33549/physiolres.933224

50. Enooku K, Kondo M, Fujiwara N, Sasako T, Shibahara J, Kado A, Okushin K, Fujinaga H, Tsutsumi T, Nakagomi R, et al. Hepatic IRS1 and ss-catenin expression is associated with histological progression and overt diabetes emergence in NAFLD patients. J Gastroenterol. 2018;53:1261–1275. doi: 10.1007/s00535-018-1472-0

51. Honma M, Sawada S, Ueno Y, Murakami K, Yamada T, Gao J, Kodama S, Izumi T, Takahashi K, Tsukita S, et al. Selective insulin resistance with differential expressions of IRS-1 and IRS-2 in human NAFLD livers. Int J Obes (Lond). 2018;42:1544–1555. doi: 10.1038/s41366-018-0062-9

52. Liss KH, Finck BN. PPARs and nonalcoholic fatty liver disease. Biochimie. 2017;136:65–74. doi: 10.1016/j.biochi.2016.11.009

53. Montagner A, Polizzi A, Fouche E, Ducheix S, Lippi Y, Lasserre F, Barquissau V, Regnier M, Lukowicz C, Benhamed F, et al. Liver PPARalpha is crucial for whole-body fatty acid homeostasis and is protective against NAFLD. Gut. 2016;65:1202–1214. doi: 10.1136/gutjnl-2015-310798

54. Ferre P, Foufelle F. Hepatic steatosis: a role for de novo lipogenesis and the transcription factor SREBP-1c. Diabetes Obes Metab. 2010;12 Suppl 2:83–92. doi: 10.1111/j.1463-1326.2010.01275.x

55. Pettinelli P, Videla LA. Up-regulation of PPAR-gamma mRNA expression in the liver of obese patients: an additional reinforcing lipogenic mechanism to SREBP-1c induction. J Clin Endocrinol Metab. 2011;96:1424–1430. doi: 10.1210/jc.2010-2129

56. Moreno-Vedia J, Girona J, Ibarretxe D, Masana L, Rodriguez-Calvo R. Unveiling the Role of the Fatty Acid Binding Protein 4 in the Metabolic-Associated Fatty Liver Disease. Biomedicines. 2022;10. doi: 10.3390/biomedicines10010197

57. Wood GC, Chu X, Argyropoulos G, Benotti P, Rolston D, Mirshahi T, Petrick A, Gabrielson J, Carey DJ, DiStefano JK, et al. A multi-component classifier for nonalcoholic fatty liver disease (NAFLD) based on genomic, proteomic, and phenomic data domains. Scientific reports. 2017;7:43238. doi: 10.1038/srep43238

58. Sveinbjornsson G, Ulfarsson MO, Thorolfsdottir RB, Jonsson BA, Einarsson E, Gunnlaugsson G, Rognvaldsson S, Arnar DO, Baldvinsson M, Bjarnason RG, et al. Multiomics study of nonalcoholic fatty liver disease. Nature genetics. 2022;54:1652–1663. doi: 10.1038/s41588-022-01199-5

59. Gu J, Liu S, Du S, Zhang Q, Xiao J, Dong Q, Xin Y. Diagnostic value of MRI-PDFF for hepatic steatosis in patients with non-alcoholic fatty liver disease: a meta-analysis. European radiology. 2019;29:3564–3573. doi: 10.1007/s00330-019-06072-4

60. Bril F, Ortiz-Lopez C, Lomonaco R, Orsak B, Freckleton M, Chintapalli K, Hardies J, Lai S, Solano F, Tio F, Cusi K. Clinical value of liver ultrasound for the diagnosis of nonalcoholic fatty liver disease in overweight and obese patients. Liver Int. 2015;35:2139–2146. doi: 10.1111/liv.12840

61. Perakakis N, Stefanakis K, Mantzoros CS. The role of omics in the pathophysiology, diagnosis and treatment of non-alcoholic fatty liver disease. Metabolism. 2020;111S:154320. doi: 10.1016/j.metabol.2020.154320

62. Kotronen A, Peltonen M, Hakkarainen A, Sevastianova K, Bergholm R, Johansson LM, Lundbom N, Rissanen A, Ridderstrale M, Groop L, et al. Prediction of non-alcoholic fatty liver disease and liver fat using metabolic and genetic factors. Gastroenterology. 2009;137:865–872. doi: 10.1053/j.gastro.2009.06.005

63. Walker RW, Belbin GM, Sorokin EP, Van Vleck T, Wojcik GL, Moscati A, Gignoux CR, Cho J, Abul-Husn NS, Nadkarni G, et al. A common variant in PNPLA3 is associated with age at diagnosis of NAFLD in patients from a multi-ethnic biobank. Journal of hepatology. 2020;72:1070–1081. doi: 10.1016/j.jhep.2020.01.029

64. Corbin KD, Abdelmalek MF,Spencer MD, da Costa KA, Galanko JA, Sha W, Suzuki A, Guy CD, Cardona DM, Torquati A, et al. Genetic signatures in choline and 1-carbon metabolism are associated with the severity of hepatic steatosis. FASEB journal: official publication of the Federation of American Societies for Experimental Biology. 2013;27:1674–1689. doi: 10.1096/fj.12-219097

65. Fairfield CJ, Drake TM, Pius R, Bretherick AD, Campbell A, Clark DW, Fallowfield JA, Hayward C, Henderson NC, Joshi PK, et al. Genome-Wide Association Study of NAFLD Using Electronic Health Records. Hepatol Commun. 2022;6:297–308. doi: 10.1002/hep4.1805

66. Anstee QM, Darlay R, Cockell S, Meroni M, Govaere O, Tiniakos D, Burt AD, Bedossa P, Palmer J, Liu YL, et al. Genome-wide association study of non-alcoholic fatty liver and steatohepatitis in a histologically characterised cohort(⋆). Journal of hepatology. 2020;73:505–515. doi: 10.1016/j.jhep.2020.04.003

67. Tan Y, Ge G, Pan T, Wen D, Gan J. A pilot study of serum microRNAs panel as potential biomarkers for diagnosis of nonalcoholic fatty liver disease. PLoS One. 2014;9:e105192. doi: 10.1371/journal.pone.0105192

68. Wu J, Zhang R, Shen F, Yang R, Zhou D, Cao H, Chen G, Pan Q, Fan J. Altered DNA Methylation Sites in Peripheral Blood Leukocytes from Patients with Simple Steatosis and Nonalcoholic Steatohepatitis (NASH). Med Sci Monit. 2018;24:6946–6967. doi: 10.12659/MSM.909747

69. Dessein A. Clinical utility of polygenic risk scores for predicting NAFLD disorders. Journal of hepatology. 2021;74:769–770. doi: 10.1016/j.jhep.2021.02.005

70. Oh HS, Rutledge J, Nachun D, Palovics R, Abiose O, Moran-Losada P, Channappa D, Urey DY, Kim K, Sung YJ, et al. Organ aging signatures in the plasma proteome track health and disease. Nature. 2023;624:164–172. doi: 10.1038/s41586-023-06802-1

71. Deczkowska A, David E, Ramadori P, Pfister D, Safran M, Li B, Giladi A, Jaitin DA, Barboy O, Cohen M, et al. XCR1(+) type 1 conventional dendritic cells drive liver pathology in non-alcoholic steatohepatitis. Nat Med. 2021;27:1043–1054. doi: 10.1038/s41591-021-01344-3

72. Rennert K, Steinborn S, Groger M, Ungerbock B, Jank AM, Ehgartner J, Nietzsche S, Dinger J, Kiehntopf M, Funke H, et al. A microfluidically perfused three dimensional human liver model. Biomaterials. 2015;71:119–131. doi: 10.1016/j.biomaterials.2015.08.043

73. Hassan S, Sebastian S, Maharjan S, Lesha A, Carpenter AM, Liu X, Xie X, Livermore C, Zhang YS, Zarrinpar A. Liver-on-a-Chip Models of Fatty Liver Disease. Hepatology. 2020;71:733–740. doi: 10.1002/hep.31106

74. Du K, Li S, Li C, Li P, Miao C, Luo T, Qiu B, Ding W. Modeling nonalcoholic fatty liver disease on a liver lobule chip with dual blood supply. Acta Biomater. 2021;134:228–239. doi: 10.1016/j.actbio.2021.07.013

75. Levner D, Ewart L. Integrating Liver-Chip data into pharmaceutical decision-making processes. Expert Opin Drug Discov. 2023;18:1313–1320. doi: 10.1080/17460441.2023.2255127

76. Ewart L, Apostolou A, Briggs SA, Carman CV, Chaff JT, Heng AR, Jadalannagari S, Janardhanan J, Jang KJ, Joshipura SR, et al. Performance assessment and economic analysis of a human Liver-Chip for predictive toxicology. Commun Med (Lond). 2022;2:154. doi: 10.1038/s43856-022-00209-1

77. Jang KJ, Otieno MA, Ronxhi J, Lim HK, Ewart L, Kodella KR, Petropolis DB, Kulkarni G, Rubins JE, Conegliano D, et al. Reproducing human and cross-species drug toxicities using a Liver-Chip. Sci Transl Med. 2019;11. doi: 10.1126/scitranslmed.aax5516

78. Cong Y, Han X, Wang Y, Chen Z, Lu Y, Liu T, Wu Z, Jin Y, Luo Y, Zhang X. Drug Toxicity Evaluation Based on Organ-on-a-chip Technology: A Review. Micromachines (Basel). 2020;11. doi: 10.3390/mi11040381

79. Gori M, Simonelli MC, Giannitelli SM, Businaro L, Trombetta M, Rainer A. Investigating Nonalcoholic Fatty Liver Disease in a Liver-on-a-Chip Microfluidic Device. PLoS One. 2016;11:e0159729. doi: 10.1371/journal.pone.0159729

80. Sun BB, Chiou J, Traylor M, Benner C, Hsu YH, Richardson TG, Surendran P, Mahajan A, Robins C, Vasquez-Grinnell SG, et al. Plasma proteomic associations with genetics and health in the UK Biobank. Nature. 2023;622:329–338. doi: 10.1038/s41586-023-06592-6

81. Wagenknecht LE, Perkins LL, Cutter GR, Sidney S, Burke GL, Manolio TA, Jacobs DR, Jr., Liu KA, Friedman GD, Hughes GH, et al. Cigarette smoking behavior is strongly related to educational status: the CARDIA study. Preventive medicine. 1990;19:158–169.

82. Dyer AR, Cutter GR, Liu KQ, Armstrong MA, Friedman GD, Hughes GH, Dolce JJ, Raczynski J, Burke G, Manolio T. Alcohol intake and blood pressure in young adults: the CARDIA Study. Journal of clinical epidemiology. 1990;43:1–13.

83. Bild DE, Jacobs DR, Jr., Sidney S, Haskell WL, Anderssen N, Oberman A. Physical activity in young black and white women. The CARDIA Study. Ann Epidemiol. 1993;3:636–644.

84. Sidney S, Jacobs DR, Jr., Haskell WL, Armstrong MA, Dimicco A, Oberman A, Savage PJ, Slattery ML, Sternfeld B, Van Horn L. Comparison of two methods of assessing physical activity in the Coronary Artery Risk Development in Young Adults (CARDIA) Study. Am J Epidemiol. 1991;133:1231–1245.

85. Jiao J, Watt GP, Lee M, Rahbar MH, Vatcheva KP, Pan JJ, McCormick JB, Fisher-Hoch SP, Fallon MB, Beretta L. Cirrhosis and Advanced Fibrosis in Hispanics in Texas: The Dominant Contribution of Central Obesity. PLoS One. 2016;11:e0150978. doi: 10.1371/journal.pone.0150978

86. de Ledinghen V, Vergniol J, Capdepont M, Chermak F, Hiriart JB, Cassinotto C, Merrouche W, Foucher J, Brigitte le B. Controlled attenuation parameter (CAP) for the diagnosis of steatosis: a prospective study of 5323 examinations. J Hepatol. 2014;60:1026–1031. doi: 10.1016/j.jhep.2013.12.018

87. Wilman HR, Kelly M, Garratt S, Matthews PM, Milanesi M, Herlihy A, Gyngell M, Neubauer S, Bell JD, Banerjee R, Thomas EL. Characterisation of liver fat in the UK Biobank cohort. PLoS One. 2017;12:e0172921. doi: 10.1371/journal.pone.0172921

88. Watt GP, De La Cerda I, Pan JJ, Fallon MB, Beretta L, Loomba R, Lee M, McCormick JB, Fisher-Hoch SP. Elevated Glycated Hemoglobin Is Associated With Liver Fibrosis, as Assessed by Elastography, in a Population-Based Study of Mexican Americans. Hepatol Commun. 2020;4:1793–1801. doi: 10.1002/hep4.1603

89. van der Poorten D, Samer CF, Ramezani-Moghadam M, Coulter S, Kacevska M, Schrijnders D, Wu LE, McLeod D, Bugianesi E, Komuta M, et al. Hepatic fat loss in advanced nonalcoholic steatohepatitis: are alterations in serum adiponectin the cause? Hepatology. 2013;57:2180–2188. doi: 10.1002/hep.26072

90. Storey JD, Tibshirani R. Statistical significance for genomewide studies. Proc Natl Acad Sci U S A. 2003;100:9440–9445. doi: 10.1073/pnas.1530509100

91. Robinson MD, Oshlack A. A scaling normalization method for differential expression analysis of RNA-seq data. Genome Biol. 2010;11:R25. doi: 10.1186/gb-2010-11-3-r25

92. Chatterjee E, Rodosthenous RS, Kujala V, Gokulnath P, Spanos M, Lehmann HI, de Oliveira GP, Shi M, Miller-Fleming TW, Li G, et al. Circulating extracellular vesicles in human cardiorenal syndrome promote renal injury in a kidney-on-chip system. JCI Insight. 2023;8. doi: 10.1172/jci.insight.165172

93. Yu G, Wang LG, Han Y, He QY. clusterProfiler: an R package for comparing biological themes among gene clusters. OMICS. 2012;16:284–287. doi: 10.1089/omi.2011.0118

94. Szklarczyk D, Kirsch R, Koutrouli M, Nastou K, Mehryary F, Hachilif R, Gable AL, Fang T, Doncheva NT, Pyysalo S, et al. The STRING database in 2023: protein-protein association networks and functional enrichment analyses for any sequenced genome of interest. Nucleic Acids Res. 2023;51:D638–D646. doi: 10.1093/nar/gkac1000

95. Shannon P, Markiel A, Ozier O, Baliga NS, Wang JT, Ramage D, Amin N, Schwikowski B, Ideker T. Cytoscape: a software environment for integrated models of biomolecular interaction networks. Genome Res. 2003;13:2498–2504. doi: 10.1101/gr.1239303

96. Jain A, Tuteja G. TissueEnrich: Tissue-specific gene enrichment analysis. Bioinformatics. 2019;35:1966–1967. doi: 10.1093/bioinformatics/bty890

97. Uhlen M, Fagerberg L, Hallstrom BM, Lindskog C, Oksvold P, Mardinoglu A, Sivertsson A, Kampf C, Sjostedt E, Asplund A, et al. Proteomics. Tissue-based map of the human proteome. Science. 2015;347:1260419. doi: 10.1126/science.1260419

98. Gonzales TI, Westgate K, Strain T, Hollidge S, Jeon J, Christensen DL, Jensen J, Wareham NJ, Brage S. Cardiorespiratory fitness assessment using risk-stratified exercise testing and dose-response relationships with disease outcomes. Sci Rep. 2021;11:15315. doi: 10.1038/s41598-021-94768-3

99. Wu P, Gifford A, Meng X, Li X, Campbell H, Varley T, Zhao J, Carroll R, Bastarache L, Denny JC, et al. Mapping ICD-10 and ICD-10-CM Codes to Phecodes: Workflow Development and Initial Evaluation. JMIR Med Inform. 2019;7:e14325. doi: 10.2196/14325

