## Supplemental Material for "A prognostic molecular signature of hepatic steatosis is spatially heterogeneous and dynamic in human liver"

Supplemental Data File

SD01 – Protein dictionary

SD02 – Regression model results for liver attenuation as a function of individual proteins adjusted for age, sex, race, and BMI in CARDIA. Note that FDR calculation in the validation sample was only performed on proteins with an FDR<5% in the derivation sample.

SD03 – Regression model results for liver attenuation as a function of individual proteins adjusted for age, sex, race, BMI, alcoholic drinks per week, systolic blood pressure, use of hypertensive and cholesterol medications, diabetes, pack years of smoking, total cholesterol, high-density lipoprotein, estimated glomerular filtration rate, and moderate-vigorous physical activity in CARDIA. Note that FDR calculation in the validation sample was only performed on proteins with an FDR<5% in the derivation sample.

SD04 – LASSO model coefficients from CARDIA for the full and top 21 protein scores of liver attenuation.

SD05 – LASSO model coefficients from the recalibrated proteins scores for UK Biobank and CCHC.

SD06 – Cox model results from UK Biobank. Full adjustment includes age, sex, race, BMI, Townsend Deprivation Index, diabetes, smoking, alcohol use, systolic blood pressure, and LDL. We conducted a sensitivity analysis including further adjustment for AST, ALT and A1c.

SD07 – Results from liver-on-a-chip experiment for hepatocytes.

SD08 – Results from liver-on-a-chip experiment for non-parenchymal cells.

**Supplemental Table 1: Baseline characteristics of CARDIA study population.** Continuous measures are reported as median (25th percentile, 75th percentile). Categorical measures are reported as n (%). Percent missingness is reported after the semi-colon for each cell. P-values are from Wilcoxon rank sum tests for continuous measures and chi square tests for categorical measures comparing the derivation and validation subsamples.

| **Characteristic** | **Overall N = 2,679** | **Derivation N = 1,876** | **Validation N = 803** | **p-value** |
|---|---|---|---|---|
| Age | 51.0 (47.0, 53.0); 0% | 50.0 (47.0, 53.0); 0% | 51.0 (47.0, 53.0); 0% | 0.2 |
| Sex | | | | 0.3 |
| Male | 1,149 (43%); 0% | 793 (42%); 0% | 356 (44%); 0% | |
| Female | 1,530 (57%); 0% | 1,083 (58%); 0% | 447 (56%); 0% | |
| Race | | | | >0.9 |
| Black | 1,259 (47%); 0% | 882 (47%); 0% | 377 (47%); 0% | |
| White | 1,420 (53%); 0% | 994 (53%); 0% | 426 (53%); 0% | |
| CARDIA field center | | | | 0.5 |
| Birmingham, AL | 632 (24%); 0% | 433 (23%); 0% | 199 (25%); 0% | |
| Chicago, IL | 623 (23%); 0% | 435 (23%); 0% | 188 (23%); 0% | |
| Minneapolis, MN | 707 (26%); 0% | 491 (26%); 0% | 216 (27%); 0% | |
| Oakland, CA | 717 (27%); 0% | 517 (28%); 0% | 200 (25%); 0% | |
| BMI ($kg/m^2$) | 29 (25, 34); 0% | 29 (25, 34); 0% | 29 (25, 34); 0% | 0.5 |
| Drinks/week | 1.0 (0.0, 5.0); 0% | 1.0 (0.0, 5.0); 0% | 1.0 (0.0, 5.0); 0% | 0.8 |
| Mean systolic blood pressure (mmHg) | 117 (108, 127); 0.1% | 117 (108, 127); 0.1% | 117 (108, 127); 0.1% | 0.6 |
| Mean diastolic blood pressure (mmHg) | 73 (66, 80); 0.1% | 73 (66, 80); 0.2% | 73 (66, 81); 0.1% | >0.9 |
| On anti-hypertensive therapy | 724 (27%); 0% | 504 (27%); 0% | 220 (27%); 0% | 0.8 |
| On cholesterol lowering medication | 438 (16%); 0% | 312 (17%); 0% | 126 (16%); 0% | 0.5 |
| Diabetes | 395 (15%); 0% | 280 (15%); 0% | 115 (14%); 0% | 0.7 |
| Lifetime pack-years smoking | 0 (0, 6); 0% | 0 (0, 6); 0% | 0 (0, 7); 0% | 0.9 |
| Total cholesterol (mg/dL) | 190 (166, 215); 0% | 190 (166, 215); 0% | 189 (167, 212); 0% | 0.9 |
| High-density lipoprotein (mg/dL) | 55 (45, 67); 0% | 54 (45, 67); 0% | 55 (44, 66); 0% | 0.6 |
| eGFR ($ml/min/1.73m^2$) | 94 (82, 108); <0.1% | 94 (82, 108); 0% | 95 (82, 109); 0.2% | 0.2 |
| Hemoglobin A1c (%) | 5.50 (5.30, 5.80); 1.2% | 5.50 (5.30, 5.80); 1.1% | 5.50 (5.30, 5.80); 1.2% | >0.9 |
| Sum of AHA Life Simple 7 | 9.00 (7.00, 10.00); 21% | 9.00 (7.00, 10.00); 21% | 9.00 (7.00, 10.00); 23% | 0.5 |
| Visceral adipose tissue volume ($cm^3$) | 118 (77, 171); 0.7% | 119 (78, 171); 0.6% | 116 (74, 174); 0.7% | 0.6 |
| Subcutaneous adipose tissue volume ($cm^3$) | 308 (212, 446); 0.7% | 310 (214, 447); 0.6% | 301 (204, 443); 0.7% | 0.2 |
| Mean liver attenuation (Hounsfield units, HU) | 58 (51, 62); 0% | 58 (51, 62); 0% | 58 (51, 62); 0% | 0.8 |
| MASLD (defined as liver attenuation <40 HU) | 268 (10%); 0% | 183 (9.8%); 0% | 85 (11%); 0% | 0.5 |

**Supplemental Table 2: UK Biobank Participants.** Continuous measures are reported a median (25th percentile, 75th percentile). Categorical measures are reported as n (%). P-values are from Wilcoxon rank sum tests for continuous measures and chi square tests for categorical measures comparing participants with and without liver MRI data.

| **Characteristic** | **Overall N = 26,421** | **Missing liver MRI data N = 24,310** | **Liver MRI data present N = 2,111** | **p-value** |
|---|---|---|---|---|
| Age | 58 (50, 64) | 59 (51, 64) | 54 (47, 60) | <0.001 |
| Female | 14,242 (54%) | 13,128 (54%) | 1,114 (53%) | 0.3 |
| Race | | | | <0.001 |
| Asian | 534 (2.0%) | 512 (2.1%) | 22 (1.0%) | |
| Black | 564 (2.1%) | 545 (2.2%) | 19 (0.9%) | |
| Mixed | 177 (0.7%) | 161 (0.7%) | 16 (0.8%) | |
| Unknown-other | 432 (1.6%) | 411 (1.7%) | 21 (1.0%) | |
| White | 24,714 (94%) | 22,681 (93%) | 2,033 (96%) | |
| Body mass index (kg/m$^2$) | 26.8 (24.2, 29.9) | 26.9 (24.3, 30.0) | 25.9 (23.6, 28.6) | <0.001 |
| Unknown | 121 | 119 | 2 | |
| Systolic blood pressure (mmHg) | 138 (125, 152) | 138 (126, 152) | 134 (122, 147) | <0.001 |
| Unknown | 1,609 | 1,446 | 163 | |
| Diabetes | 1,540 (5.8%) | 1,497 (6.2%) | 43 (2.0%) | <0.001 |
| Unknown | 28 | 28 | 0 | |
| Townsend Deprivation Index | -2.1 (-3.6, 0.7) | -2.0 (-3.6, 0.8) | -2.6 (-3.9, -0.1) | <0.001 |
| Unknown | 35 | 32 | 3 | |
| Smoking | | | | <0.001 |
| Current | 2,814 (11%) | 2,677 (11%) | 137 (6.5%) | |
| Never \| No Answer | 14,350 (54%) | 13,082 (54%) | 1,268 (60%) | |
| Previous | 9,226 (35%) | 8,520 (35%) | 706 (33%) | |
| Unknown | 31 | 31 | 0 | |
| Alcohol frequency | | | | <0.001 |
| Never \| No Answer | 2,290 (8.7%) | 2,185 (9.0%) | 105 (5.0%) | |
| Special occasions only | 3,137 (12%) | 2,956 (12%) | 181 (8.6%) | |
| One to three times a month | 2,817 (11%) | 2,615 (11%) | 202 (9.6%) | |
| Once or twice a week | 6,919 (26%) | 6,378 (26%) | 541 (26%) | |
| Three or four times a week | 5,946 (23%) | 5,348 (22%) | 598 (28%) | |
| Daily or almost daily | 5,281 (20%) | 4,797 (20%) | 484 (23%) | |
| Unknown | 31 | 31 | 0 | |
| Liver proton density fat fraction | 3.0 (2.2, 5.1) | - | 3.0 (2.2, 5.1) | |
| Unknown | 24,310 | - | 0 | |
| Aspartate aminotransferase (U/L) | 24 (21, 29) | 24 (21, 29) | 24 (21, 28) | <0.001 |
| Unknown | 1,343 | 1,239 | 104 | |
| Alanine aminotransferase (U/L) | 20 (15, 27) | 20 (15, 27) | 19 (15, 26) | <0.001 |
| Unknown | 1,261 | 1,163 | 98 | |
| Hemoglobin A1c (mmol/mol) | 35.3 (32.8, 38.1) | 35.4 (32.9, 38.2) | 34.4 (31.9, 36.8) | <0.001 |
| Unknown | 1,370 | 1,253 | 117 | |
| Low density lipoprotein (mmol/L) | 3.48 (2.91, 4.10) | 3.48 (2.90, 4.10) | 3.49 (2.93, 4.08) | 0.8 |
| Unknown | 1,321 | 1,219 | 102 | |

**Supplemental Table 3: Baseline characteristics of the Cameron County Hispanic Cohort.** Continuous measures are reported a median (25$^{th}$ percentile, 75$^{th}$ percentile). Categorical measures are reported as n (%). Hypertension defined as systolic blood pressure ≥ 140mmHg or diastolic blood pressure ≥90mmHg.

| Characteristic | N | |
|---|---|---|
| Age (years) | 206 | 56 (46, 66) |
| Female | 206 | 136 (66%) |
| White Hispanic/Latino | 206 | 206 (100%) |
| Body mass index (kg/m$^2$) | 206 | 30.9 (27.4, 35.8) |
| Type 2 Diabetes | 206 | 66 (32%) |
| Hypertension | 201 | 22 (11%) |
| Alcohol use | 128 | |
| Never | | 63 (49%) |
| Sometimes | | 52 (41%) |
| Other | | 13 (10%) |
| Controlled attenuation parameter (dB/m) | 206 | 297 (247, 337) |

**Supplementary Table 4: Cells used for liver-on-a-chip quad culture**

| Cell Names | Company | Catalogue No. | Source |
|---|---|---|---|
| Cryo Human Hepatocytes | Gibco | HU8305 | Normal human donor |
| Primary Human Liver Sinusoidal MVEC | Cell Systems | ACBRI 566 | Normal human donor |
| Human Liver Kupffer Cells | SAMSARA Science | HLKC | Normal human donor |
| Human Hepatic Stellate Cells | iXCells Biotechnologies | 10HU-210 | Normal human donor |

**Supplementary Table 5: Primer list for liver-on-a-chip experiments.**

| Sl. No. | Gene Names | Sequences (5 ⟶3) |
|---|---|---|
| 1. | HMGCS1 F | AAGTCACACAAGATGCTACACCG |
| | HMGCS1 R | TCAGCGAAGACATCTGGTGCC |
| 2. | SERPINE1 F | CTCATCAGCCACTGGAAAGGCA |
| | SERPINE1 R | GACTCGTGAAGTCAGCCTGAAA |
| 3. | HSPA1B F | ACCTTCGACGTGTCCATCCTGA |
| | HSPA1B R | TCCTCCACGAAGTGGTTCACCA |
| 4. | ENO3 F | TGGGAAGGATGCCACCAATGTG |
| | ENO3 R | GCGATAGAACTCAGATGCTGCC |
| 5. | HSPA1A F | ACCTTCGACGTGTCCATCCTGA |
| | HSPA1A R | TCCTCCACGAAGTGGTTCACCA |
| 6. | ME1 F | GGAGTTGCTCTTGGTGTTGTGG |
| | ME1 R | GGATAAAGCCGACCCTCTTCCA |
| 7. | CTSZ F | GGATGGTGTCAACTATGCCAGC |
| | CTSZ R | CACGCTCCCTTCCTCTTGATGT |
| 8. | DEFB1 F | GGTAACTTTCTCACAGGCCTTGG |
| | DEFB1 R | TCCCTCTGTAACAGGTGCCTTG |
| 8. | PYGL F | CACTTCAGTGGCAGATGTGGTG |
| | PYGL R | GCAGTGGAAATCTGCTCTGACAG |
| 9 | CDA F | GCCAAGAAGTCAGCCTACTGCC |
| | CDA R | CTTCTGGATAGCGGTCCGTTCA |
| 10. | IL1RAP F | CTGAGGATCTCAAGCGCAGCTA |
| | IL1RAP R | AGCAGGACTGTGGCTCCAAAAC |
| 11. | SHBG F | CAGGACAAGAGCCTATCGCTGT |
| | SHBG R | GTCATCCTTAGGGTTGGTATCCC |
| 12. | IRS1F | AGTCTGTCGTCCAGTAGCACCA |
| | IRS1 R | ACTGGAGCCATACTCATCCGAG |
| 13. | IRS2 F | CCTGCCCCCTGCCAACACCT |
| | IRS2 R | TGTGACATCCTGGTGATAAAGCC |
| 14. | PPARα F | TCGGCGAGGATAGTTCTGGAAG |
| | PPARα R | GACCACAGGATAAGTCACCGAG |
| 15. | PPARγ F | AGCCTGCGAAAGCCTTTTGGTG |
| | PPARγ R | GGCTTCACATTCAGCAAACCTGG |
| 16. | FABP4 F | ACGAGAGGATGATAAACTGGTGG |
| | FABP4 R | GCGAACTTCAGTCCAGGTCAA |
| 17. | SREBP1c F | ACTTCTGGAGGCATCGCAAGCA |
| | SREBP1cR | AGGTTCCAGAGGAGGCTACAAG |
| 18. | β-ACTIN F | CACCATTGGCAATGAGCGGTTC |
| | β-ACTIN R | AGGTCTTTGCGGATGTCCACGT |

**Supplemental Figure 1: Stability and validation of regression estimates in CARDIA.**
(A) There was high correlation between beta coefficients from linear models relating individual aptamers to liver attenuation adjusted age, sex, race, BMI adjusted with beta coefficients from the “full” adjusted (age, sex, race, BMI, alcoholic drinks per week, systolic blood pressure, hypertension and cholesterol medication use, diabetes, pack years, total cholesterol, high-density lipoprotein, estimated glomerular filtration rate, and moderate-vigorous physical activity) in CARDIA validate sample. See Supplemental Data File SD03 for full regression results. (B) There was moderate correlation of beta coefficients from linear models relating individual aptamers to liver attenuation adjusted age, sex, race, BMI between CARDIA derivation and validation subsamples. See Supplemental Data File SD02 for full regression results.

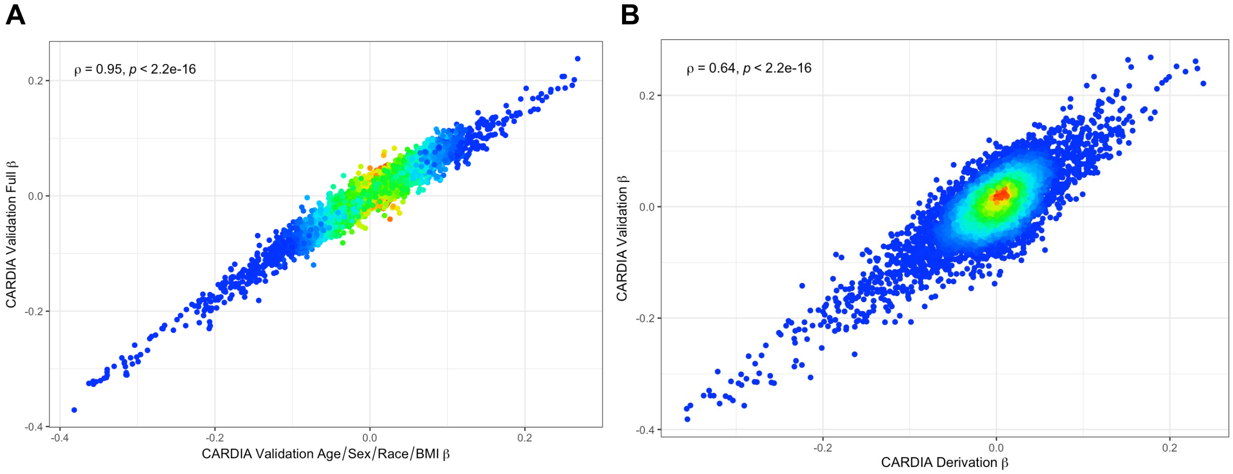

**Supplemental Figure 2: Proteomics of liver attenuation and their relationship with age, sex, race, BMI, and alcohol use in CARDIA and UK Biobank.**

(A & B) Protein score of liver attenuation is not strongly related to age. (C & D) Protein score of liver attenuation not significantly different across sex and race. (E & F) There is a weak correlation between the protein score of liver attenuation and body mass index (BMI). (G & H) Protein score of liver attenuation does not vary greatly by alcohol use.

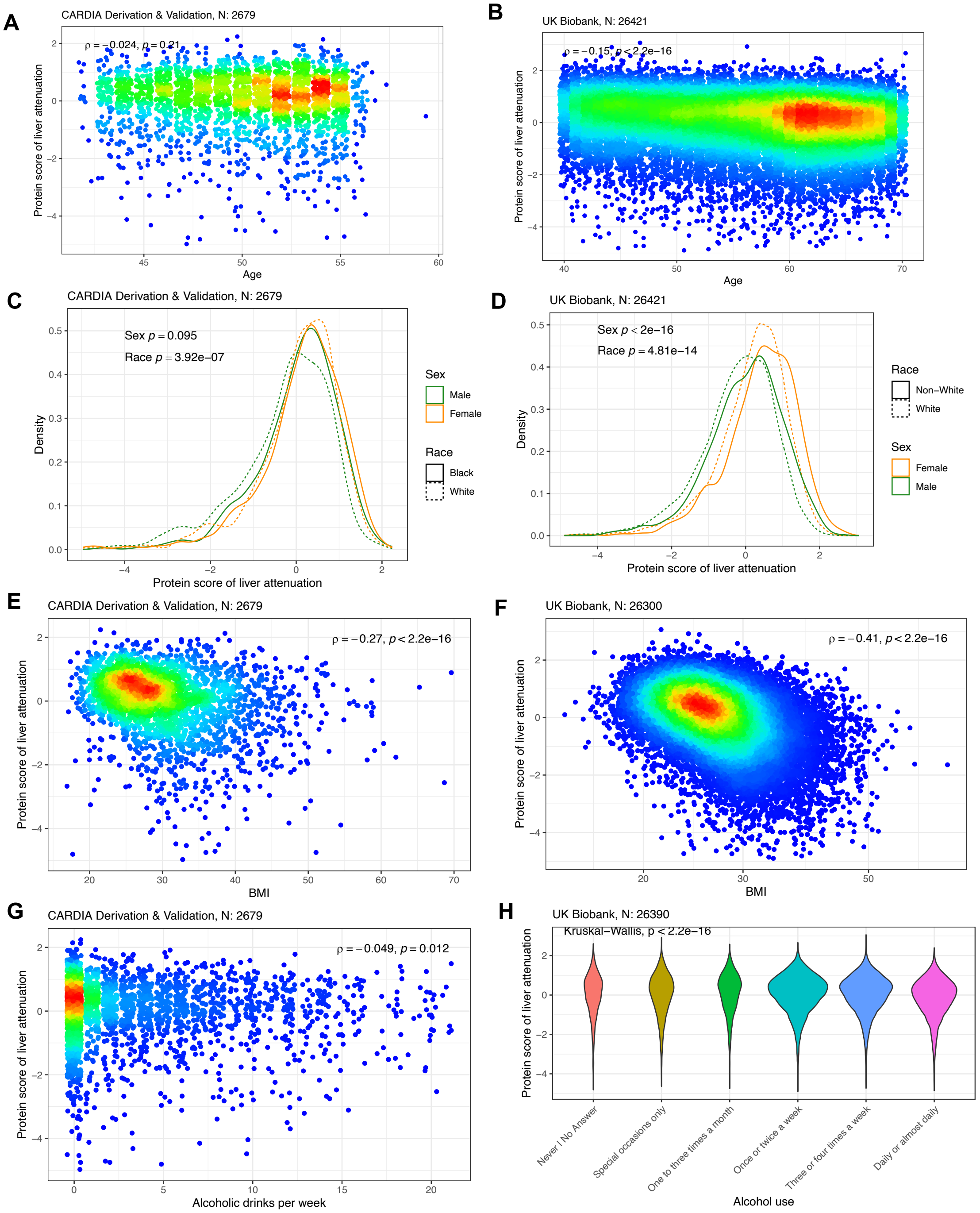

**Supplemental Figure 3: Model fit of the top 21 protein score for hepatic steatosis in CARDIA and UK Biobank.** (A) Top 21 protein score of liver attenuation by CT (less attenuation ~ more steatosis) demonstrated moderate correlation with the parent variable in both CARDIA derivation and validation samples. The top 21 proteins were defined by ranking the absolute value of the beta coefficients from the full protein score. (B) Replication of the association between top 21 protein score of liver attenuation and MRI-based measure of hepatic steatosis (proton density fat fraction: higher ~ more steatosis, opposite directionality as with CT based liver attenuation) in UK Biobank.

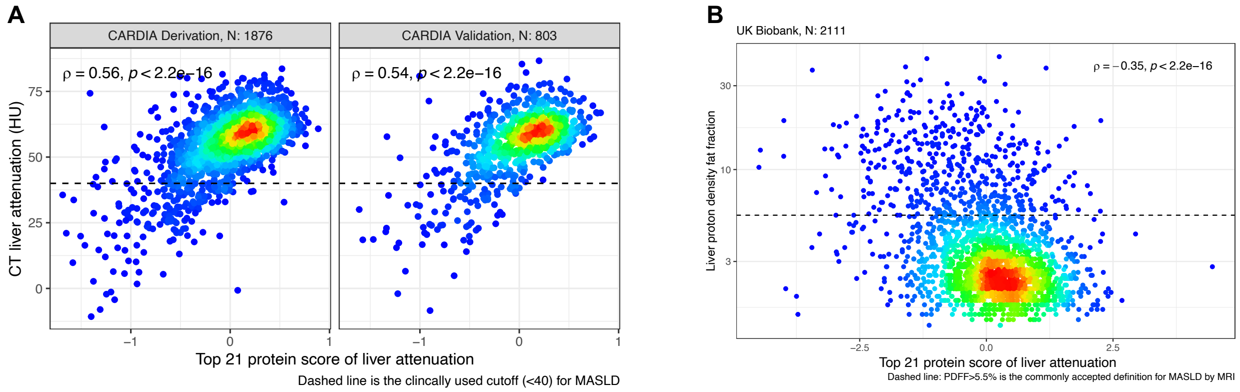

**Supplemental Figure 4: A protein score of liver attenuation is more strongly correlated with hepatic steatosis in obese CARDIA participants.** Using data from the CARDIA validation sample (N=803; N normal/overweight = 447, N obese = 356), we observed a stronger correlation between hepatic steatosis and the protein score of liver attenuation. Obese is defined as body mass index ≥ 30kg/m$^2$.

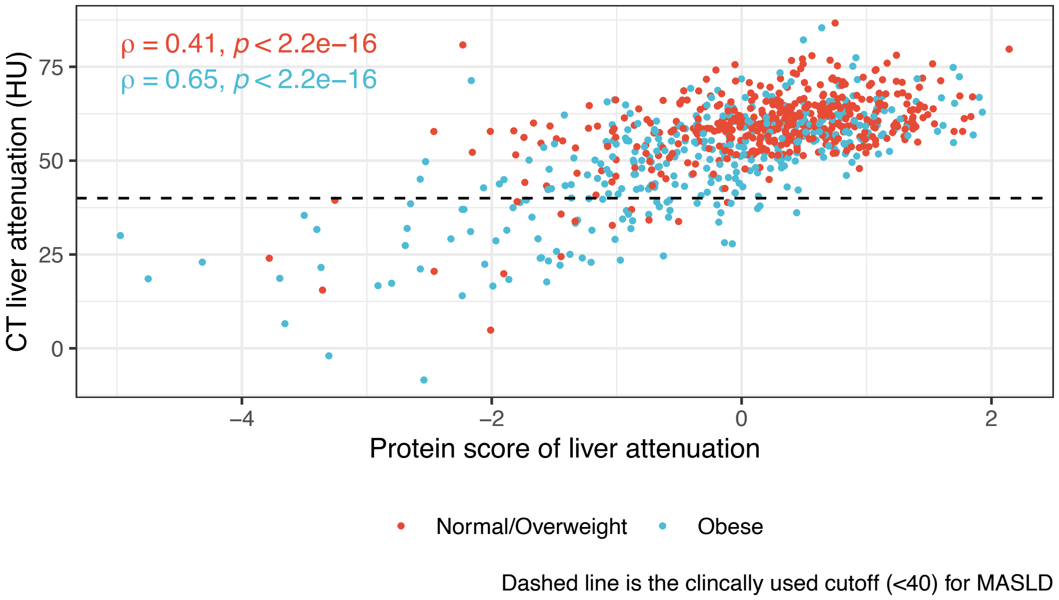

**Supplemental Figure 5 Cell-specific and spatial architecture of the MASLD proteome.** (**A**) Gene expression of 198 implicated targets by the circulating MASLD proteome (in CARDIA) and were also expressed in both scRNA-seq and Visium datasets across cell types using only the liver cell atlas scRNA-seq dataset. (**B**) Hematoxylin-eosin stained healthy and steatotic liver tissue, overlaid with composite expression score. Two examples were shown in the main text; this figure includes all sections reported in the initial work. (**C**) Hematoxylin-eosin stained healthy and steatotic liver tissues, overlaid with gene expression of the top three gene targets implicated by the CARDIA model and differentially expressed in healthy and steatotic liver tissue. (**D**) Violin plots showing the top 4 model candidate gene expression across spots from healthy and fatty liver samples (Visium) and a representative model target that was not differentially expressed using our differential expression criteria (*IGF1*).

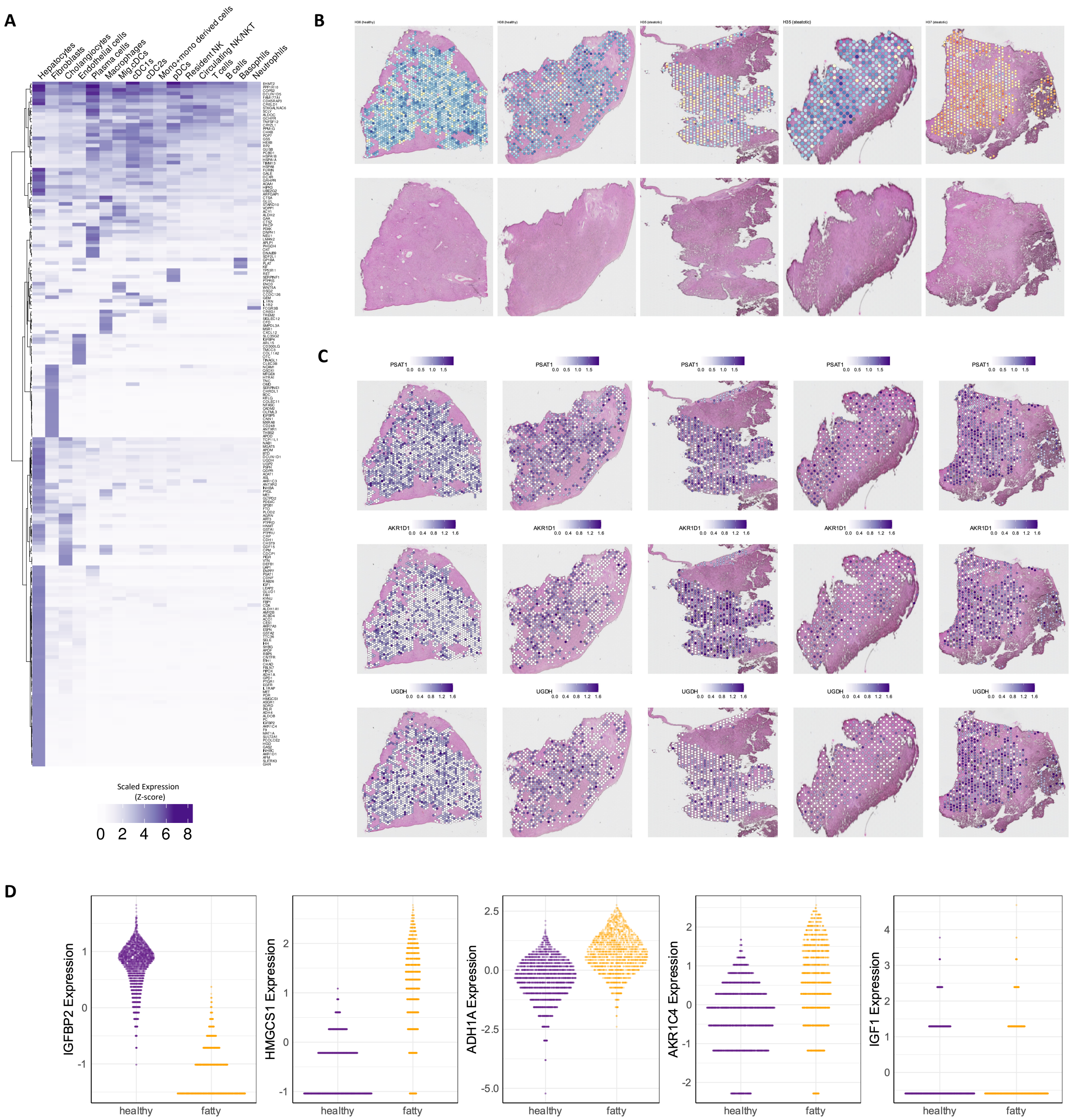

A
Hepatocytes
Fibroblasts
Cholangiocytes
Endothelial cells
Plasma cells
Macrophages
Mig.cDCs
cDC1s
cDC2s
Mono+mono derived cells
pDCs
Resident NK
Circulating NK/NKT
T cells
B cells
Basophils
Neutrophils
Scaled Expression
(Z-score)
0 2 4 6 8
B
C
PSAT1
0.0 0.5 1.0 1.5
AKR1D1
0.0 0.4 0.8 1.2 1.6
UGDH
0.0 0.4 0.8 1.2 1.6
D
IGFBP2 Expression
HMGCS1 Expression
ADH1A Expression
AKR1C4 Expression
IGF1 Expression
healthy
fatty

**Supplemental Figure 6: Comparison of log2 fold change with negative binomial mixed models in spatial transcriptomic analysis.** We compared the effect size estimates of our differential expression analysis to coefficients from negative binomial mixed models with sample as random effect. We observed high agreement between $\log_2$ fold change and the negative binomial mixed model coefficients for all 198 model targets (all points) and for the 33 differentially expressed genes (purple points). Pearson r reported.

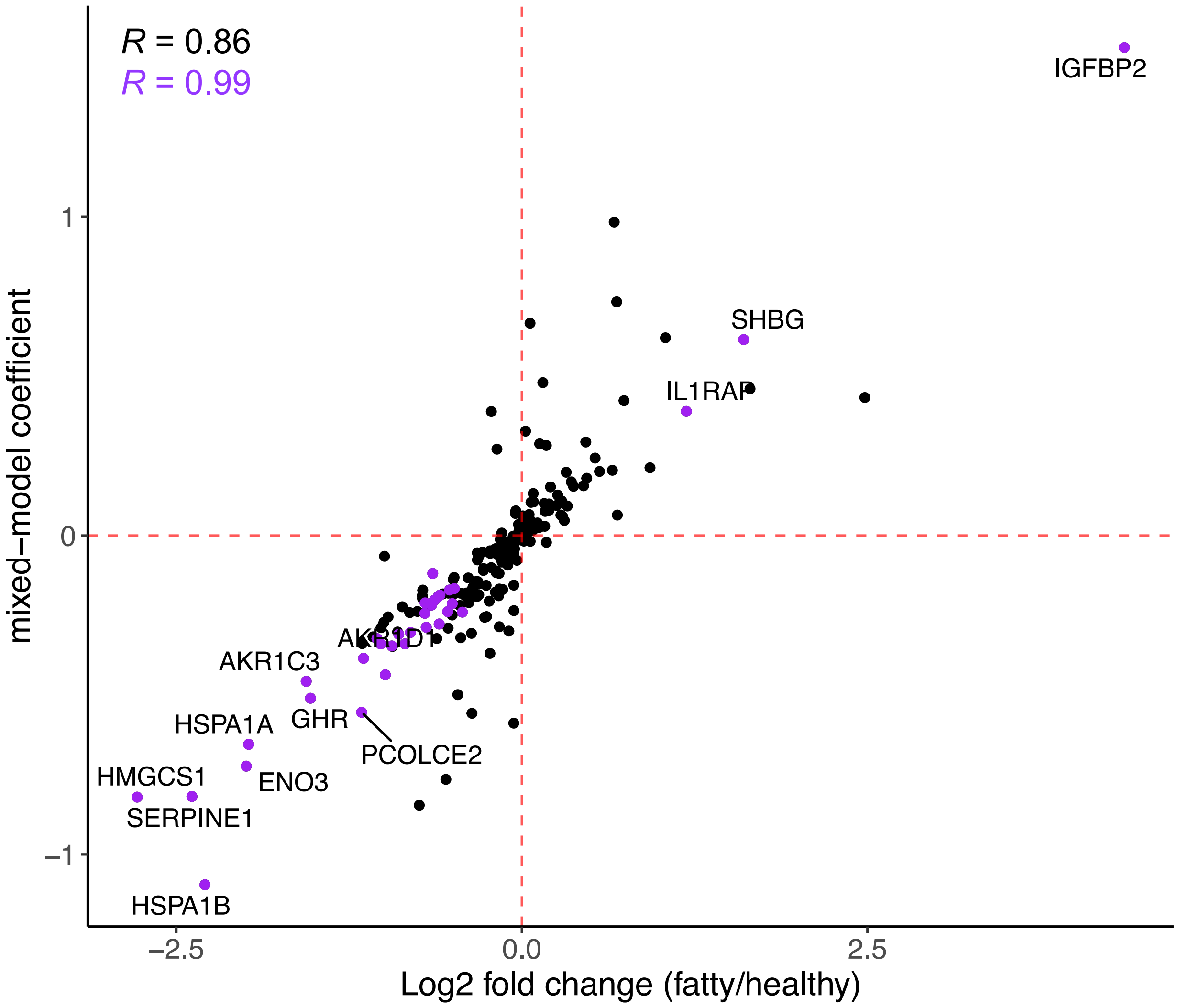
